# CAMION: a catchment area maximization algorithm, with application to oncology accessibility in metropolitan France

**DOI:** 10.1101/2021.09.29.21264296

**Authors:** Eric Daoud, Anne-Sophie Hamy, Elise Dumas, Lidia Delrieu, Beatriz Grandal Rejo, Christine Le Bihan-Benjamin, Sophie Houzard, Philippe-Jean Bousquet, Judicaël Hotton, Aude-Marie Savoye, Christelle Jouannaud, Chloé-Agathe Azencott, Marc Lelarge, Fabien Reyal

**Affiliations:** Residual Tumor & Response to Treatment Laboratory, RT2Lab, INSERM, U932 Immunity and Cancer, Institut Curie, Université Paris, 75005 Paris, France; INRIA, DI/ENS, PSL Research University, Paris, France; Department of Medical Oncology, Institut Curie, Paris, France; INSERM, U900, 75005 Paris, France; MINES ParisTech, PSL Research University, CBIO-Centre for Computational Biology, 75006 Paris, France; Department of Surgery, Institut Curie, Paris, France; Health Data and Assessment Department, Survey Data Science and Assessment Division, National Cancer Institute, 52 avenue André Morizet 92100 Boulogne-Billancourt, France; Aix Marseille Univ, Inserm, IRD, SESSTIM, Equipe Labellisée Ligue Contre le Cancer, Marseille, France; Survey Data Science and Assessment Division, National Cancer Institute, 52 avenue André Morizet 92100 Boulogne-Billancourt, France; Department of Surgery, Institut Jean Godinot, Reims, France; Department of Medical Oncology, Institut Jean Godinot, Reims, France; Institut Curie, PSL Research University, 75005 Paris, France

## Abstract

**Background:** Access to health services plays a key role in cancer survival. Uneven distributions of populations and health facilities lead to geographical disparities. Location-allocation algorithms can address these disparities by finding new locations and capacities for health facilities. However, in oncology, opening new hospitals or moving them is difficult in practice, and should be handled carefully.

**Methods:** We propose a method to measure the spatial accessibility to oncology care and identify the hospitals to grow to reduce disparities. We first ran a clustering algorithm to automatically label the hospitals in terms of oncology specialization. Then, we computed an accessibility score to these hospitals for every population location. Finally, we introduced *CAMION*, an optimization algorithm based on Linear Programming that reduces disparities in oncology accessibility by identifying health facilities that should increase their capacities.

**Results:** We demonstrate our algorithm in metropolitan France. The clustering step let us identify different oncology specialization levels for hospitals. Most of the population in metropolitan France lived in good accessibility areas, especially in large cities. Lower accessibility zones are often rural or suburban municipalities. The optimization algorithm effectively manages to identify hospitals to grow, based on current oncology specialization and accessibility scores.

**Discussion:** There is a tradeoff to be found by patients, between care center proximity and care center expertise, which is less likely to happen for patients living in good accessibility areas. The accessibility score is deliberately non-specific to cancer type but can be adapted to more precise pathologies. Our method is replicable in any country, given hospitals and population locations data. We developed a web application intended for healthcare professionals to let them to run the optimization algorithm with the desired parameters and visualize the results.

## Introduction

Cancer is a leading cause of death worldwide, accounting for nearly 10 million deaths in 2020. While a lot of the ongoing research is focusing on finding new cancer treatments, accessibility to oncology care receives less attention. Yet, several studies have showed that access to health services plays a key role in cancer survival. For instance, geographic residency status and social environment seem to explain treatment and prognosis disparities for patients with non-small cell lung cancer (1). In France, increases in travel times to health services were associated with lower survival rates for patients with a colorectal cancer (2). In New Zealand, living in deprived areas, far from a cancer center or from primary care was associated with lower survival chances for patients with colorectal, lung and prostate cancers (3).

Accessibility refers to the relative ease by which services can be reached from a given location (4). Accessibility can be defined by spatial factors, determined by where you are; and non-spatial factors, determined by who you are (5). Spatial accessibility methods assess the availability of supply locations from demand locations, connected by a travel impedance metric. Supply locations are characterized by their capacity or quantity of available resource. Similarly, demand locations are characterized by their population. Such methods have been successfully used to measure access to healthcare, such as primary care (6) or oncology care (4,7,8) in several countries including France (9–11). In what follows, we restrict accessibility to spatial accessibility and use both terms interchangeably.

Uneven distributions of population and health-care providers lead to geographic disparity in accessibility for patients (12). For instance, Weiss et al. (13) showed that 8.9% of the global population could not reach healthcare within one hour if they have access to motorized transport. In Germany, Bauer et al. (14) shown that 10% of the population lived in areas with low accessibility for internal medicine and surgery. Location-allocation algorithms (15) can optimize the distribution and supply of health providers to reduce accessibility disparities. These algorithms seek the optimal placement of facilities for a desirable objective under certain constraints (4). For instance, Luo et al. developed an optimization algorithm to improve the healthcare planning in rural China by finding the best place and capacity for new health facilities (16). Tao et al. worked on a spatial optimization model to maximize equity in accessibility to residential care facility in Beijing, China (17). When optimizing health accessibility, there are two competing goals: equity and efficiency (18,19). Equity may be defined as equal access to healthcare for everyone (20). An efficient situation is when everything has been done to help any person without harming anyone else (21). While some argue that efficiency should be addressed in priority (21), others agree that equity is a matter of ethical obligation, especially in public health (22,23).

The goal of this paper is to apply spatial accessibility methods to oncology care centers and propose an optimization algorithm to reduce disparities. We demonstrate our results in metropolitan France. There are many care centers in France, which do not share the same degree of oncology specialization. Therefore, we first run a clustering algorithm to automatically group the care centers based on their medical statistics and attributes. Using these clusters, we label the care centers in terms of hospital development and oncology specialization. Then, we compute an oncology accessibility score for every municipality in metropolitan France. We then introduce *CAMION*, an optimization algorithm based on *Linear Programming* which uses the clusters of care centers and the accessibility scores to suggest, given a limited budget, where to increase hospital capacity to improve the oncology accessibility. Finally, our method is packaged into a web application intended to healthcare professionals so they can run the optimization algorithm with the desired parameters for any region.

## Methods

### Data collection

Health data is collected from two sources: the *French national administrative database* (PMSI) and the *French annual health facilities statistics* (*SAE*). *PMSI* data includes discharge summaries for all inpatients admitted to public and private hospitals in France. The *SAE* database is a compulsory and exhaustive administrative survey of all public and private hospitals in France. The survey is sent every year and describes the activities of the hospitals as well as the list of services and their staff. We restricted the analysis to the year 2018. We in-clude every hospital in metropolitan France that declared a *Medicine, Surgery or Obstetric (MCO)* activity in the *SAE* survey, in 2018. We also included the liberal radiotherapy care centers, with no *MCO* activity. The resulting dataset contains 1,662 care centers.

Geographic and travel data were retrieved from open data platforms. Municipalities and their census statistics were extracted from the *National Statistics Bureau of France (INSEE)* website. We used the *OpenRouteService (ORS) API* to compute the driving routes between hospitals and municipalities, which is necessary for the accessibility score.

### Care centers characterization

We selected a list of 24 variables with the help of medical experts to characterize the care centers. The list of variables and their definitions is available in the supplementary materials. The variables are either binary when they encode the presence or absence of a service; or discrete when they encode the number of stays. We only focus on treatments received in hospitals.

Given the large number of care centers, we use a clustering algorithm to automatically group together similar care centers. More specifically, we first run a *Principal Component Analysis (PCA)* algorithm on the *SAE* dataset that describes the care centers. The input data has 24 variables, and we perform the dimensionality reduction with *n* = 2 components. We tried different number of components, from 2 to 5, but we found 2 gave good and easy to interpret results. We then run a clustering algorithm on the PCA-reduced dataset to automatically isolate care centers with similar statistics. We tried several algorithms like *K-Means (24), DBSCAN (25) and Spectral Clustering* (26). In our case, *Spectral Clustering* with 8 clusters gave the most interpretable and better isolated groups. For the number *k* of clusters, we tested all values from 2 to 10 and manually interpreted the results with medical experts.

### Accessibility score

There are several ways to compute accessibility to healthcare (6). The easiest and most straightforward methods are computed within bordered areas, like provider-to-population ratios in each municipality. While they are very intuitive, these methods do not account for border crossing, or travel impedance, which makes them less accurate. Recently, a new type of method has been developed and is now used in most spatial accessibility papers. This algorithm is called *Two Step Floating Catchment Area (2SFCA)* (27). It is a two-step method that first computes a provider-to-population ratio for each provider location. In the second step, for each population location, an accessibility score is obtained by summing the provider-to-population ratios. For the algorithm to work, a catchment threshold (distance or travel time) must be set. Above this threshold, a provider location is considered unreachable from the population location, and vice versa. The *2SFCA* method does not account for distance decay: a care center is either reachable or not. The *Enhanced Two Step Floating Catchment Area (e2SFCA)* (28) addresses this limitation by applying weights to differentiate travel zones in both steps.

We now explain more formally how to compute *eS2FCA* scores. Consider *P*_*i*_ the population at location *i*, with 1 ≤ *i* ≤ *n* where *n* is the number of population locations. Similarly, consider *S*_*u*_ the capacity of care center *u*, with 1 ≤ *u* ≤ *m* where *m* is the number of care centers. Finally, let *d*_*iu*_ be the matrix of size *n* × *m* containing the distances between location *i* and care center *u*. We consider *r* sub-catchment zones each associated with a weight *W*_*s*_, and a distance *D*_*s*_, with 1 ≤ *s* ≤ *r*, such that *D*_1_ < *D*_2_ < … < *D*_*r*_ and *W*_1_ > *W*_2_> … > *W*_*r*_. The resulting *r* travel intervals are *I*_1_ = [0, *D*_1_], *I*_2_= [*D*_1_, *D*_2_],…, *I*_*r*_ = [*D*_*r*−1_ − *D*_*r*_], The accessibility *A*_*i*_ of a population location *i* is computed in two steps. Step 1: for every care center *u*, compute its weighted capacity-to-population ratio *R*_*u*_. Step 2: for every population location, compute *A*_*i*_ as the sum all the weighted *R*_*u*_ of the reachable care centers.

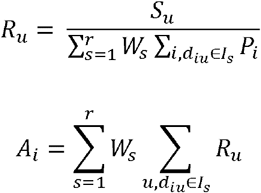

The capacity of a care center is balanced by the total population with access to it. A population location that solely has access to low capacities or overcrowded care centers will have a low accessibility score. Similarly, a population location will have low accessibility scores if the distance to get to the nearby care centers is large.

As we want to compute the accessibility to oncology care centers, we chose *S*_*u*_ to be the oncology activity of a hospital *u*. We define oncology activity as the sum of the number of medical and surgery stays related to cancer, and the number of patients with chemotherapy or radiotherapy. A care center with no oncology activity will have *R*_*u*_ = 0 and a municipality that solely has access to this care center *u* will have *A*_*i*_ = 0. We use driving duration as travel impedance metric, and we set the maximum catchment area to a 90-minute drive. In 2018, only 24,152 patients out of 761,057 (3.2%) had travel duration greater than 90 minutes for cancer related pathways. This is low enough to consider that care centers are non-reachable beyond this distance. We divide the catchment area into 3 intervals: *I*_1_ = (0, 30], *I*_2_ = (30, 60] and *I*_3_= (60, 90]. The associated weights are respectively *W*_1_ = 1, *W*_2_ = 0.042 and *W*_3_ = 0.09. These sub catchment areas are set based on the cancer pathways travel duration distributions and validated with medical experts. The weights are the same than the *e2SFCA* paper (28).

For privacy reasons, municipalities with small populations are grouped in entities called “geographic codes” in the *PMSI* data. We decided to compute the accessibility score for each geographic code and municipalities that are grouped in the same code will have the same accessibility score.

### Accessibility optimization

Regarding efficiency optimization, the most popular algorithms are *p-median*, location set covering problem *(LSCP)* and *maximum covering location problem (MCLP)*. The *p-median* algorithm minimizes the weighted sum of distances between users and facilities (29). *LSCP* minimizes the number of facilities needed to cover all demand (30). *MCLP* maximizes the demand covered within a desired distance or time threshold by locating a given number of facilities (31).

To reach equal access to healthcare, quadratic programming has been used to minimize the variance of accessibility scores defined by the *2SFCA* (32). Similarly, a *Particle Swarm Optimization (PSO)* algorithm was developed to minimize the total square difference between the accessibility score of each demand location and the weighted average accessibility score (17). Finally, a two-step optimization algorithm has been developed to address the dual objectives of efficiency and equality, by first choosing where to site new hospitals and then deciding which capacity they should have (16,33).

However, most of the previous algorithms seek locations to open new health facilities. In this work, we are interested in the case where the health facilities are fixed, and the only lever to improve accessibility is to increase their capacities. Given a capacity budget, we want to know which facilities to grow and by how much. We introduce *CAMION*, an accessibility optimization algorithm based on *Floating Catchment Area* and *Linear Programming*. The initial accessibility score was computed with the *Enhanced Two Step Floating Catchment Area (e2SFCA)* (28) but our algorithm can generalize to more *FCA* derivatives.

We model the problem as an optimization task. In our case, we want our optimization algorithm to find new care centers capacities given some constraints, so that the total accessibility is maximum. We apply optimization on a given region only, rather than on the whole metropolitan France. We chose this approach because healthcare planning is handled regionally rather than nationally. We show below that our optimization problem is a Linear Programming problem.

In its standard form, *Linear Programming* finds a vector *x* that maximizes *c*^*T*^*x* under constraints *Ax* ≤ *b*, where *A* is a matrix and *b* a vector. Boundaries can be set to *x* such as *x* ≥ 0. Consider *x*_*u*_ the new capacity of a care center *u*, to be computed by the algorithm. Let *Q*_*u*_ and *W*_*u*_ be two vectors of size *m*, defined as follows:

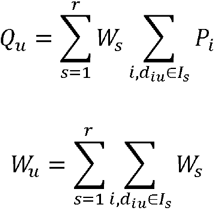

We can compute the total accessibility as a sum on the *m* care centers:

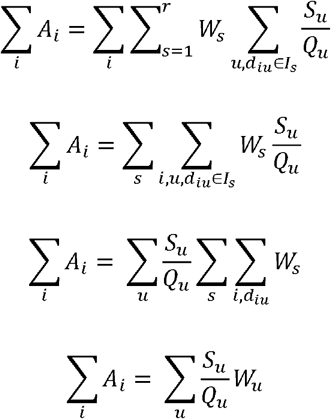

The last equation can be rewritten in the *Linear Programming* standard form with:

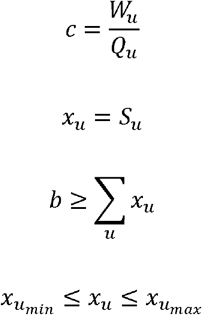

The user-defined parameters are *b*, 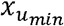 and 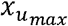. *b* is the total capacity to be shared across all the care centers. 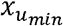 and 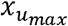 are the capacity boundaries for care center *u*. If *b* is set to the current total capacity, a care center can’t be grown unless another one is decreased. If *b* > Σ_*u*_ *x*_*u*_, the capacity of care centers can be increased without decreasing other centers. We know how to solve *Linear Programming* and we used the *SciPy* (34) implementation of the revised simplex method as explained in (35).

We now detail how we set the user-defined parameters to apply the *Linear Programming* algorithm to our specific case. The additional capacity was set as +3% of the overall activity of the region’s care centers: *b* = 1.03 × Σ_*u*_ *x*_*u*_. The choice of the boundaries 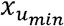 and 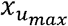 is crucial and must be realistic. We studied the hospitals activity on the past four years (2016 to 2019) to retrieve the average growth percentage of a care center. The growth percentage is computed as follows: (*S*_2019_ − *S*_2016_)/*S*_2016_. Among the care centers that grew and who had an existing oncology activity, the mean growth percentage was 23%. Hence, we set 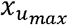 as +20% of the care center capacity. Regarding 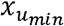, we set the boundary based on the cluster of the care center. For the three most specialized clusters, we set their 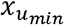 equal to their current activity. We did this to prevent the algorithm from decreasing the most specialized and well-equipped care centers. Regarding the care centers from the other clusters, 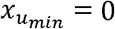, so that they could be emptied if need be. Finally, we set 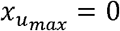 if the care center belongs to the least specialized cluster. The new capacities are indicative and should be further investigated to make sure they are relevant. Especially when setting an existing oncology activity to 0.

We developed a web application that allows the users to run the optimization algorithm in any region with the parameters they want. The application displays accessibility results and optimization outcomes on an interactive map with additional plots. The user can browse the list of care centers by cluster and the list of municipalities with their accessibility scores.

## Results

### Population and hospitals distribution in metropolitan France

In 2018, the population in France was 66,993 million. Mainland France hosts 64,812 million inhabitants (96.8%), while the remaining 2,181 million (3.2%) live in overseas departments and regions. Metropolitan France is divided into 13 administrative regions and 96 departments. The population density in France is unevenly distributed. In 2020, the overall population density in metropolitan France was 119 inhabitants per square kilometer. *Ile-de-France* region has the highest population density with 1,022 inhabitants per square kilometer. Density in other regions in metropolitan France range between 40 and 187 inhabitants/km^2^. Denser areas are located near the coastline and around the largest cities like *Paris, Marseille, Lyon, Strasbourg, Toulouse, or Bordeaux*. The middle of the country is rural, and the population densities are low. While there are a great variety of regions and landscapes, the country is becoming more urbanized. This “*rural exodus*” is largely responsible of what is known as the “*empty diagonal*”, a band of very low-density population that stretches from the southwest to the northeast.

We now describe the spatial distribution and specificities of the 1,662 hospitals included in this study. There are different types of hospitals in France: *Centres Hospitaliers* (CH, n=667) and *Centres Hospitaliers Régionaux / Universitaires* (CHR/U, n=142) are state-run hospitals; *Centres de Lutte Contre le Cancer* (CLCC, n=26) and *Participants au Service Public Hospitalier / Etablissement à But Non Lucratif* (PSPH/EBNL, n=142) are both private hospitals of collective interest, though *CLCC* are oncology dedicated; private hospitals (n=606) are privately run and for-profit. The non-*MCO* care centers with radiotherapy activity (n=79) are mostly private practice structures and are referred as *Other*. **Table 1** shows the number of care centers and their oncology activity per hospital type and region. Most of the care centers are public, but a non-neglectable part are private. *CLCC* represent only 1.6% of the care centers, yet they are responsible for 14.2% of the overall oncology activity. The care centers are unevenly distributed across the country. For instance, *Corse and Centre-Val-de-Loire* are the only two regions with no *CLCC* care centers. Moreover, the proportion of oncology activity per hospital type varies from a region to another. For instance, in *Nouvelle-Aquitaine*, 47.1% of the oncology activity is handled by private care centers, whereas in *Provence-Alpes-Cote-d’Azur* it is 21.4%.

**Table 1:**
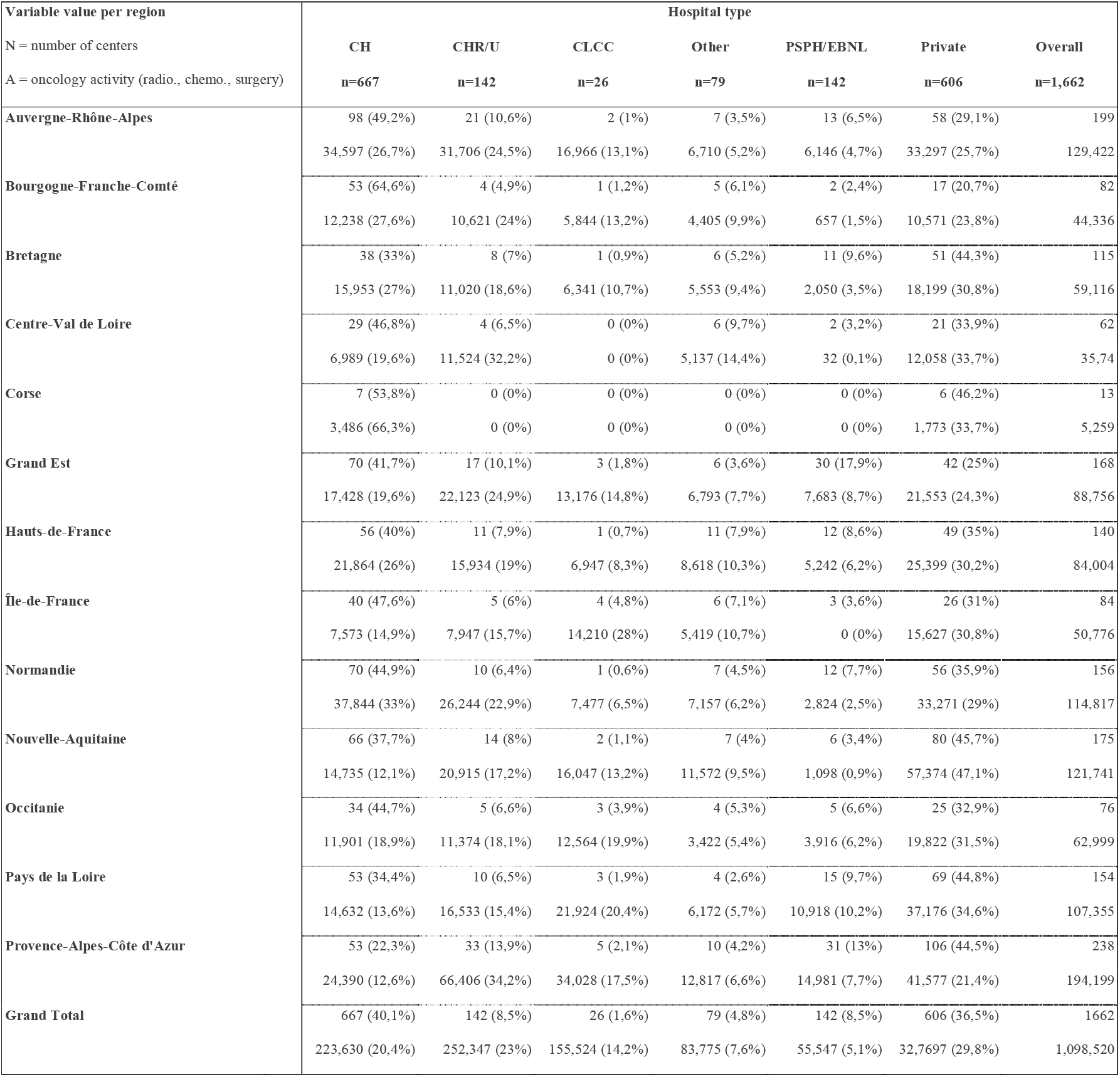
Number of care centers (N) and overall oncology activity (A) per hospital type and region. Oncology activity is the sum of the number of patients with radiotherapy or chemotherapy, and the number of medical or surgery stays related to cancer. *CH* and *CHR/U* are public hospitals; *CLCC* and *PSPH/EBNL* are private hospitals of collective interest, though *CLCC* are oncology dedicated; private hospitals are for-profit. *Other* hospitals are mostly private practice *radiotherapy* structures. The percentages sum to 100% row-wise. In *Nouvelle-Aquitaine* region, 47.1% of the oncology activity is handled by private care centers, whereas in *Provence-Alpes-Cote-d’Azur* region it is 21.4%.

### Care centers characterization

While it is obvious that *CLCC* care centers are suited for oncology care, it is difficult to assess the degree of oncology specialization for other care centers. Our clustering algorithm assigns the n=1,662 care centers into 8 clusters, sorted by oncology specialization. **Figure 1** shows the distribution of some of the key health services per cluster. These services are *biology, radiotherapy, chemotherapy, cancer surgery, intensive unit, palliative care, oncology unit, medication circuit, surgery, and outpatient surgery*. The three oncology services are *cancer surgery, radiotherapy, and chemotherapy*. We see that care centers from clusters 1 (n=79) and 2 (n=39) all have these 3 services, hence they are the most suited hospitals for oncology care. Centers from cluster 3 (n=451) have *cancer surgery* and *chemotherapy* but lack *radiotherapy*. The most part of the n=381 centers from cluster 4 have *cancer surgery*, but no *radiotherapy* nor *chemotherapy*. Care centers from cluster 5 (n=2) and cluster 6 (n=7) have *radiotherapy* and *chemotherapy* services, but no *cancer surgery*. Care centers in cluster 7 (n=77) are dedicated to *radiotherapy* and mostly private practice structures. Finally, care centers 8 (n=626) have none of the 3 oncology services. To sum up, hospitals from clusters 1 and 2 (n=118) are “all-in-one” care centers that provide the most “ideal” oncology care. Centers from clusters 3 and 4 (n=382) provide oncology care but will have to be coordinated with additional structures during the pathways. Hospitals within clusters 5, 6 and 7 (n=86) are not allowed to perform *cancer surgery* but provide *chemotherapy* or *radiotherapy*. The remaining n=626 care centers in cluster 8 are not equipped for oncology care. Hospital types are unevenly distributed among the clusters. For instance, 76.9% of the *CLCC* care centers are placed in cluster 1, as they are the most specialized centers. In cluster 7, we find external *radiotherapy* units of some CLCC centers, and private practice structures. The proportion of private care centers varies as well: cluster 1 has almost no private care center while cluster 2 has 61.5% of private hospitals. Moreover, most of the oncology activity is handled by care centers from clusters 1 and 3. Also, the overall oncology activity from the n=79 centers in cluster 1 is almost as large as the activity of the n=451 hospitals from cluster 4.

**Figure 1:**
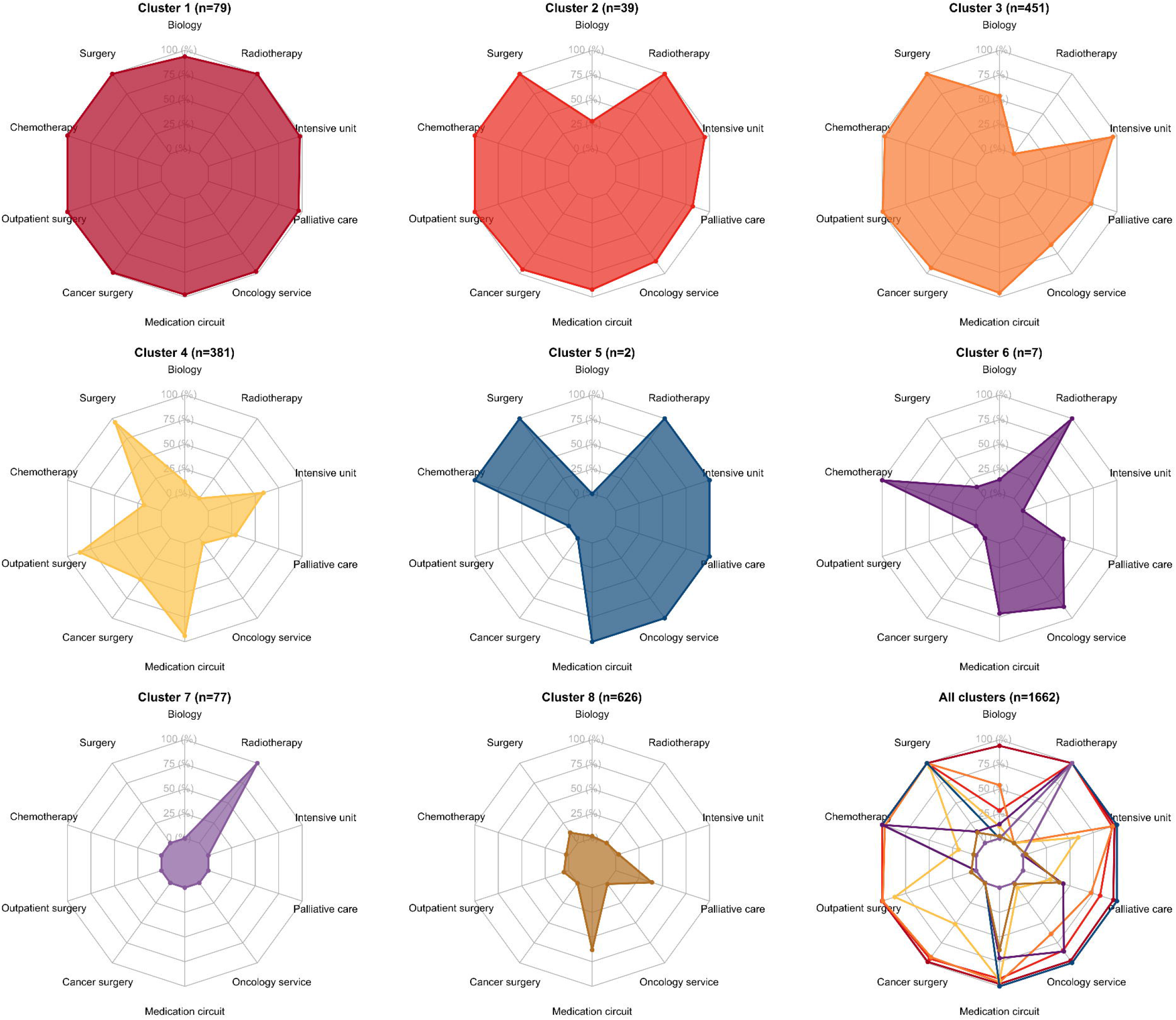
Distribution of the care centers services and equipment per cluster. Each radar plot axis shows the percentage of the care centers within the cluster that have the corresponding attribute. In Cluster 1, the care centers have all the listed services. In cluster 8, the centers have almost none of the services. Care centers from cluster 1 (n=79) and cluster 2 (n=39) are the most suited for oncology care.

### Accessibility score computation

We computed the spatial accessibility score to these care centers for every municipality in metropolitan France, using the *e2SFCA* algorithm and oncology activity as supply variable. We compared the accessibility distributions with *e2SFCA* vs. regular *2SFCA*. The accessibility was lower with *e2SFCA* because of the weight decay. We also studied the influence of the supply variable in the accessibility score. Accessibility is much higher if we use the number of *Medical, Surgery and Obstetric* (*MCO*) stays as supply, instead of the oncology activity. This makes sense since oncology care centers are less common and the overall *MCO* activity is higher than the oncology activity. The oncology accessibility is unevenly distributed across the country, as displayed on **Figure 2**. For better readability, we cut the accessibility scores into 5 quantiles. *Q5* colored in dark green contains the top 20% accessibility municipalities, and *Q1* in light yellow contains the bottom 20% ones. The lowest accessibility zones are mostly located in the center of the country and in mountainous regions like the *Alps* or the *Pyrenees*. Plot (B) shows that most of the population (51.6%) lives in top 20% accessibility municipalities, while 6.3 % lives in the bottom 20% quantile. On map (A), care centers are displayed as squares, colored by cluster index, and sized by oncology activity. We see that accessibility is highest near the most specialized care centers. Indeed, the proportion of care centers from specialized clusters decreases in lower accessibility quantiles (C). We then ranked the departments by median accessibility and showed the top-10 and bottom-10 on plot (D). Among the top-5 departments, 4 are in *Ile-de-France*. Departments from the bottom-10 are rural or mountainous areas like *Lozère* and *Alpes-de-Haute-Provence*. We notice disparities within departments as well, as outlined by the large interquartile range in *Hérault* or *Alpes-Maritimes*. On the contrary, this spread is very narrow in *Ile-de-France* departments.

**Figure 2:**
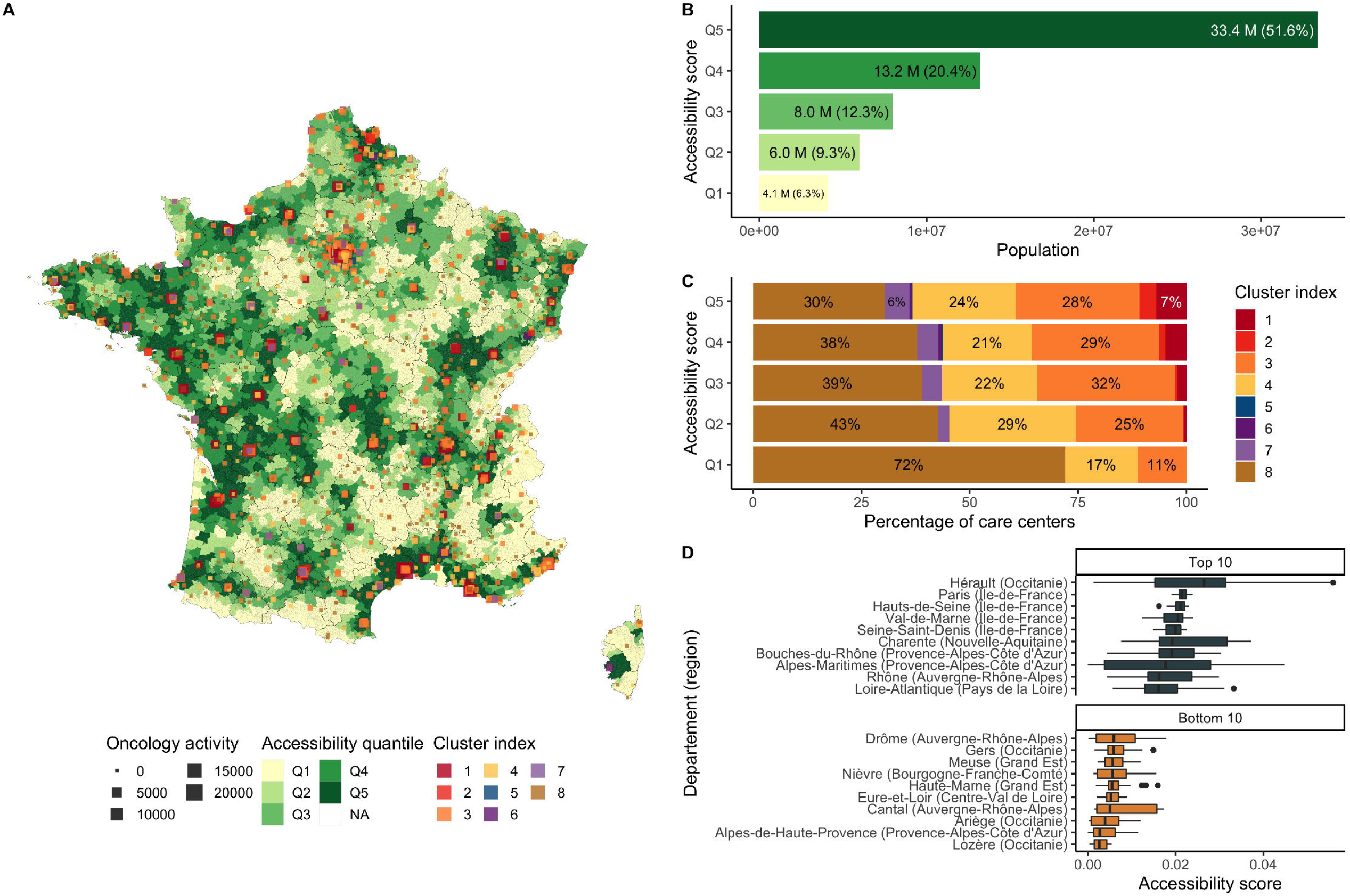
Distribution of the accessibility score computed with enhanced two step floating catchment area (e2SFCA), in metropolitan France. Plot (A) shows municipalities colored by accessibility quantile. The care centers are drawn as squares, colored by cluster, and sized by oncology activity. Plot (B) shows the total population by accessibility quantile. Plot (C) displays the percentage of care centers by cluster by accessibility quantile. Plot (D) shows the top 10 and bottom 10 list of the departments, ranked by median accessibility.

Accessibility score should be put into perspective with population density. Overall, the denser municipalities have a median accessibility around 0.02. Municipalities with low population densities have more extreme values. **Figure 3** compares accessibility and population density for three different regions: *Provence-Alpes-Cote-d’Azur* (A), *Ile-de-France* (B), and *Bourgogne-Franche-Comté* (C). Municipalities are displayed as squares, colored by accessibility quantile, and sized by population density. These regions show very different profiles. In *Provence-Alpes-Cote-d’Azur* (A), accessibility is essentially low in non-dense municipalities near the *Alps*. However, in *Bourgogne-Franche-Comté* (C), we see dense municipalities with poor accessibility scores, representing a large proportion of the region. We also drew similar maps (D, E and F) where municipalities are colored based on the average travel duration for patients with cancer in 2018. We see that the average travel time is higher in municipalities with poor accessibility scores. The surface percentage with low accessibility varies from a region to another. For instance, in *Bourgogne-Franche-Comté*, 34.5% of the region has a Q1 accessibility, that is 15.6% of the region’s population. Sometimes, the Q1 surface can be large but might contain very few inhabitants. This happens in *Ile-de-France*, where 15% of the surface is Q1 accessibility, representing less than 1% of the region’s population. Finally, we compared our accessibility score with the department exit ratio, by municipality. Department exit ratio is defined as the proportion of cancer patients who visited a care center outside from their department of residence and was computed using the *PMSI* database. In *Provence-Alpes-Cote-d’Azur*, the exit ratio is higher in departments with low accessibility scores and few oncology specialized care centers, as in *Alpes-de-Haute-Provence* and *Hautes-Alpes*. While the *Var* department has some oncology centers, exit ratio remains high since larger care centers are in *Marseille* and *Nice*.

**Figure 3:**
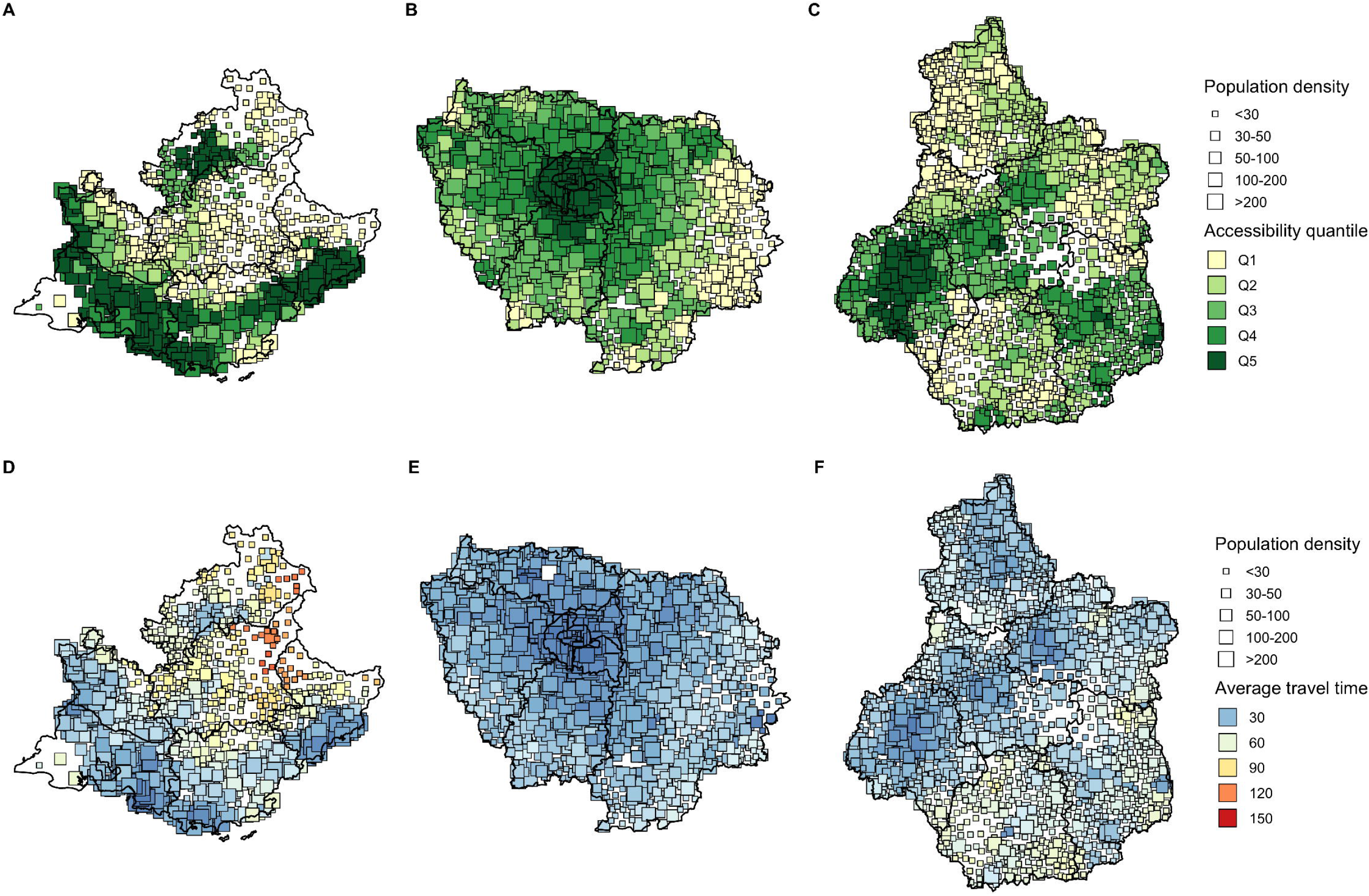
Comparison of population density with accessibility scores and patient average travel time for cancer pathways. Showing results in three regions: *Provence-Alpes-Cote-d’Azur* (A, D), *Ile-de-France* (B, E) and *Bourgogne-Franche-Comté* (C, F). Municipalities are drawn as squares, sized by population density and colored by either accessibility quantile (A, B, C) or patient average travel time (D, E, F).

We now focus on the region *Provence-Alpes-Cote-d’Azur*. This region is the far southeastern on the mainland. The region’s population was 5,048 million in 2018. Its prefecture and largest city is *Marseille*. The region contains six departments. *Bouches-du-Rhone, Var* and *Alpes-Maritimes* are located on the coastline and gather the largest cities like *Marseille, Nice*, or *Toulon. Alpes-de-Haute-Provence, Vaucluse, and Hautes-Alpes* are inland departments, with a majority of rural and mountainous areas. Results are shown on **Figure 4**. By comparing maps (A) and (B), we confirm that the accessibility is maximum in denser areas of the region. Average patients travel time are displayed on map (C) and we drew the major roads (primary, motorway, and truck) in red. The road system is well developed on the coast, rallying the larger cities of the region. However, driving from the rural areas in the *Alps* to the major cities is hard, resulting in higher travel times. The accessibility is unevenly spread within the departments, especially in *Alpes-Maritimes* where the distribution is multi-modal (D). There, cities like *Nice* and *Cannes* have large hospitals thus good accessibility, while the northern areas of the department are mostly mountains. Accessibility is higher in municipalities with dense populations, for all the departments (E). Finally, the average travel time decreases when the accessibility score increases. This makes sense since the accessibility score was computed based on the driving distance between population locations and care centers. However, it confirms that patients living in poor accessibility zones effectively travel further to seek oncology care. In *Bouches-du-Rhone*, nearly all the municipalities have an average travel time lower than 30 minutes, while in *Alpes-de-Haute-Provence*, average travel times are rarely lower than 60 minutes (F).

**Figure 4:**
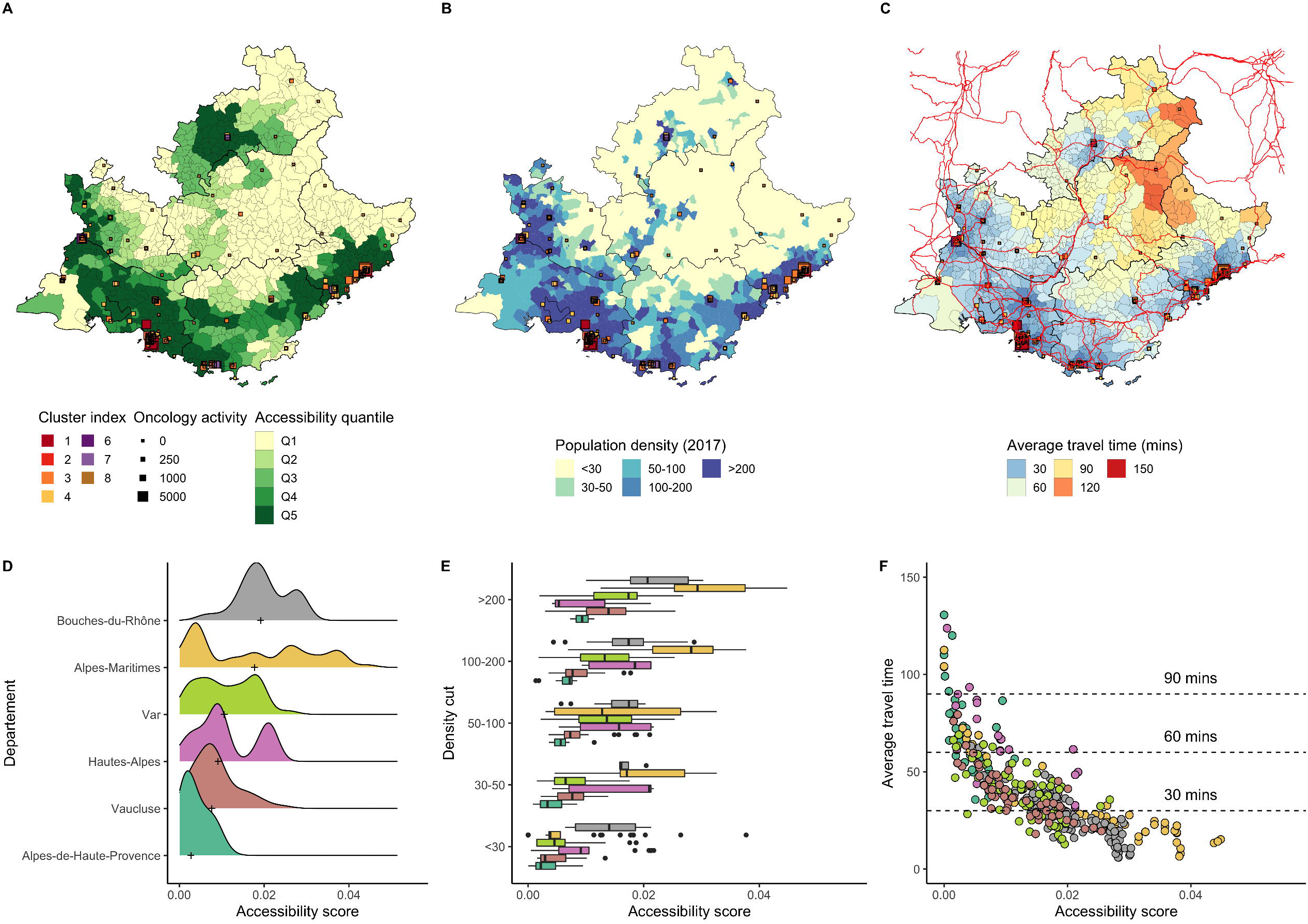
Accessibility distribution in Provence-Alpes-Cote-d’Azur region. Map (A) shows the region accessibility distribution per municipality. Map (B) displays the population density discretized in 5 bins. The map on plot (C) displays the average travel time for cancer pathways. Large roads (primary, motorway, and trucks) are drawn in red. Plot (D) shows the accessibility distribution per department of the region. Plot (E) shows the accessibility distribution by municipality population density and department. Plot (F) compares the accessibility score from municipalities with the average travel time for cancer pathways.

### Accessibility optimization

Since we focused on describing the accessibility situation in *Provence-Alpes-Cote-d’Azur*, we now present the outcomes of our optimization algorithm in this same region. The algorithm was run with the user-specified parameters stated in the Methods Section: we chose to increase the overall oncology activity in the region by 3% (+3,221 activity) and capped care centers to a 20% maximum growth. The median accessibility in the region went from 0.0093 to 0.0103, a 11.1% increase. The results are shown on **Figure 5**. Map (A) displays the accessibility delta 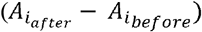 as well as the care centers eligible to grow. Centers from cluster 8 were hidden since we considered that they couldn’t provide any oncology activity. The algorithm identified a list of 26 care centers where the oncology activity could grow to maximize the total accessibility in the region. These centers are either public or private hospitals, primarily located in the *Avignon* and *Gap* areas. The care centers located in high accessibility areas near *Marseille* and *Nice* were ignored by the algorithm because improving these zones is not a priority. The care center that grew the most is *Clinique Sainte Catherine*, in *Avignon*. Interestingly, this care center was recently bought by the *Unicancer* group, which coordinates all the cancer centers in France. This hospital’s type will change to become a new *CLCC*. Thus, it is expected to grow in the next years and to be equipped with more oncology services and staff.

**Figure 5:**
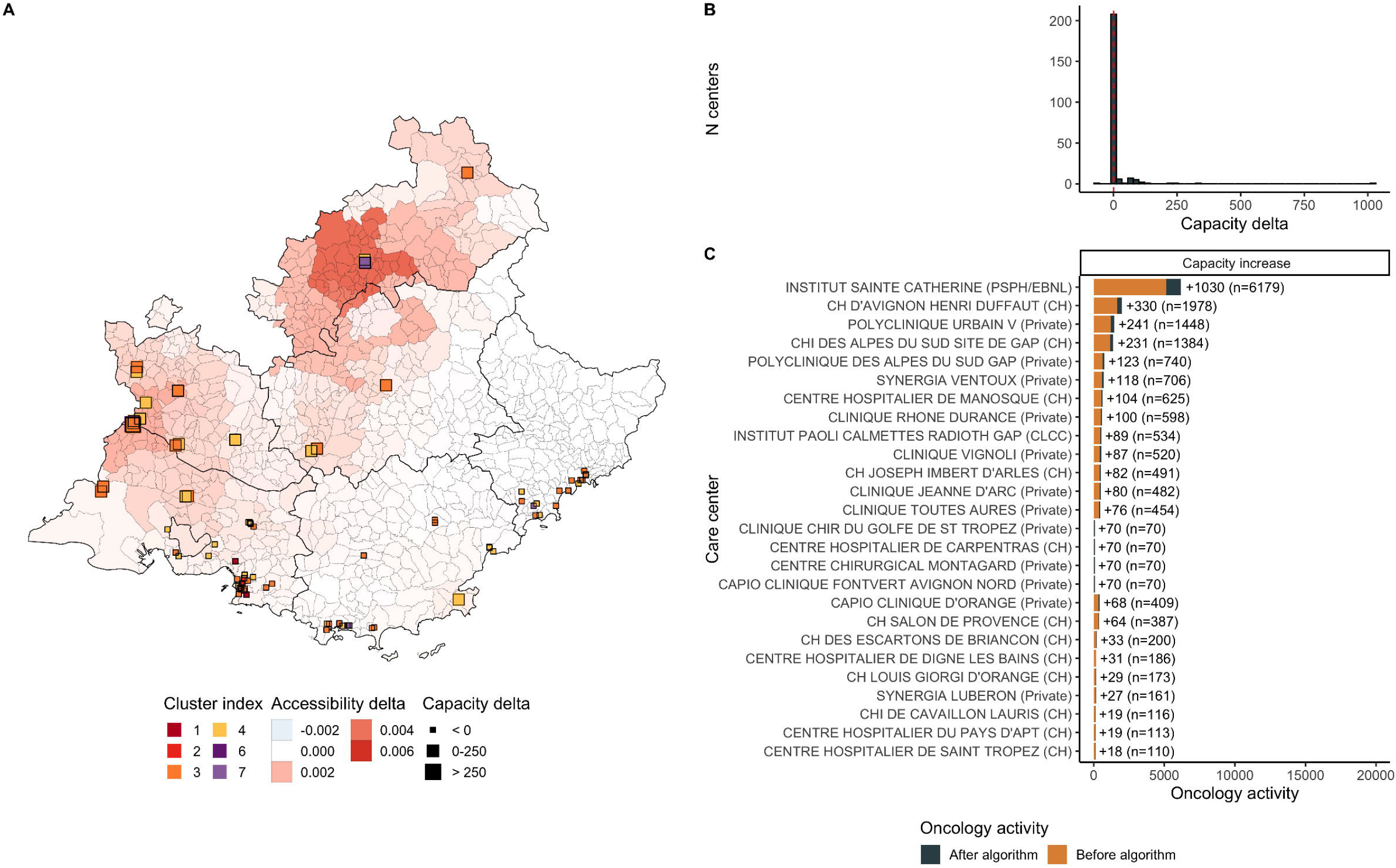
Accessibility delta in Provence-Alpes-Cote-d’Azur (PACA) region after running the optimization algorithm. Map (A) displays the accessibility delta 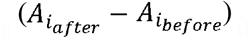 by municipality. Plot (B) shows the capacity delta 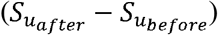 distribution. Capacity was defined as the oncology activity: the number of patients with chemotherapy or radiotherapy and the number of medical or surgery stays related to oncology. We show the list of the care centers that grew the most (C) and by how much. For instance, the hospital *Institut Sainte Catherine* in *Avignon*, was assigned a +1,030 capacity, for a total of n=6,179. Additional activity was 3,221. 26 centers grew and 1 decreased. Median accessibility before optimization was 0.0093 and 0.0103 after, corresponding to a 11.1% increase. Accessibility increased around cities like *Avignon* and *Gap*. Care centers near *Nice* were left unchanged by the algorithm.

While we described the results in *Provence-Alpes-Cote-d’Azur* region, we ran the algorithm with similar parameters on every region in metropolitan France. The results are available in the *Supplementary Materials* and on the web application. We observe two types of optimization strategies. For most regions, the algorithm manages to find a couple of areas where the accessibility can be locally improved, like it did in *Provence-Alpes-Cote-d’Azur* near *Gap* and *Avignon*. However, for regions like *Ile-de-France* and *Haut-de-France*, the hospital capacity increase is more uniformly distributed across the region. Most of the time, the algorithm left untouched the large care centers located in dense cities with good accessibilities. This can be explained by the relatively low value of the additional activity parameter: with a very large value of additional activity, every care center will grow. If we keep it low, the algorithm identifies in which areas hospital capacity should be increased in priority.

## Discussion

We observe disparities in both care centers and their accessibility. The clustering algorithm successfully groups similar hospitals and lets us identify the care centers best suited for oncology care. Some variables in the *SAE* survey are declarative and potentially differ from the reality. We are aware of this bias, but we do not expect major differences that could distort our clustering results.

Receiving treatment in a care center with *surgery, chemotherapy* and *radiotherapy* activities is easier for the patient and leads to better care pathways. Care centers from cluster 1 will be the better choice for cancer treatment and correspond to modern oncology care specifications. However, these centers are a minority and sparsely located, essentially in dense areas and in large cities. While the inhabitants of large cities and metropolitan areas will have no problem reaching them, rural areas residents live far away from these centers. This population often has better access to care centers from intermediate clusters. Such centers do not have all the key services and the patients are more likely to visit multiple hospitals during their care pathways.

Longer drives to reach a more specialized care center could be considered more acceptable for *surgery*, where the hospital volumetry and surgeon expertise matter. However, for more frequent interventions like *chemotherapy* and *radiotherapy* especially, patients should prioritize short travels. There is a tradeoff to be found by patients, between care center proximity and care center expertise. This dilemma will be more frequent for patients living in rural areas than patients living in dense cities with large care centers nearby.

Specific attention should be given to municipalities with very poor access to oncology care centers. While we saw that most of the population lives in high accessibility areas, around 6% of the population lives in the bottom 20% accessibility quantile. Among these municipalities, some are very rural and mountainous like those in the *Alpes-de-Haute-Provence* in *Provence-Alpes-Cote-d’Azur* region. Such areas cannot be expected to have a very good healthcare coverage. By contrast, the case of suburban areas with relatively dense population and poor accessibility should be addressed more easily. Our optimization algorithm can help driving public health policies, as it effectively identifies areas where accessibility could grow, by allocating additional oncology activity to a restricted number of care centers. The proposed growth factors are indicative and do not have to be effective within a year, as it represents a considerable effort for care centers to increase their activity.

Our oncology accessibility score is deliberately non-specific to cancer type. This score is meant to outline how easy it would be for a population location to reach a first entry point for oncology care. Here, we are only focusing on surgery, chemotherapy, and radiotherapy treatments. The same technique could be used on a specific cancer type, the method will remain the same, only the supply variable used in the accessibility score will change. We should mention that spatial accessibility is better suited for pathologies that are relatively well handled across the whole country. Accessibility for rare diseases like pediatric cancer or complex cancers that require a specific expertise is less informative because only a handful of care centers are indicated.

Similarly, we could compute an accessibility score that is focused on specific kinds of stays: our web application lets the user pick between surgery, chemotherapy, or radiotherapy as supply variable.

The quality of oncology care is linked with the care centers’ volumetry. A care center with a very low activity is less likely to provide decent care. As a result, the *French National Institute of Cancer (INCa)* defined several thresholds (36) that forbid care centers with very low activity to keep operating. Similarly, the care quality in a saturated care center won’t be good either, since patients are more likely to wait longer before diagnosis or between interventions. While it is easy to spot care centers with low activity, it is harder to judge if a care center is over-crowded, and we should be careful when attributing new activity to the hospitals. We based the 20% max growth out of the previous centers’ activity increase. This percentage could be tailored to the center cluster or current activity. Volumetry is not the only factor determining care quality. More sophisticated indicators like average delay between diagnosis and first treatment can tell whether a care center is in line with the care pathways recommendations. Care centers with activities lower than the thresholds, or with a large proportion of degraded pathways should be handled with care by our algorithm.

Accessibility optimization depends on many factors and healthcare professionals will not have the same uses for our algorithm. Some may consider that for a care center to grow another should decline, where others would rather not decrease any centers’ activities. Moreover, the healthcare planning is very different from a region to another, and even within the regions departments are showing disparities. Hence, we cannot expect the algorithm to be used with the same parameters on every region. For all these reasons, we believe that providing a web application to run the algorithm and choose the parameters is the most useful way to the help healthcare professionals improve the current situation.

Our work is in line with the *French Cancer Plan* (37) that emphasizes the importance of increasing accessibility to oncology care as well as minimizing disparities across the country. The government mandated *INCa* to work on the accessibility development. This study and the web application we developed could help when attributing the care centers authorizations. Working closely with researchers from *INCa* and public health professionals could have a major impact on the oncology care spatial organization in metropolitan France, benefiting millions of patients.

We ran this method in metropolitan France, but it could work on any country if data on hospitals and municipalities are available.

## Data Availability

Data available on request from the authors.

## Acknowledgements

The authors thank Thomas Ansart and the *Sciences Po* cartography workshop for helping us to improve our figures. We also thank Julien Guerin and Johan Archinard, from the *Data Factory at Institut Curie*, who helped us to deploy our web application. Finally, we thank Hakim Idjis, Marc-Felix Degni and Olivier Auliard and their team at *Capgemini Invent*, who helped us to extract the driving routes with OpenRouteService.

