## Supplementary figures for "CAMION: a catchment area maximization algorithm, with application to oncology accessibility in metropolitan France"

### Supplementary Materials

| **SAE table** | **Variable name** | **Variable definition** | **Distribution** |
| --- | --- | --- | --- |
| FILTRE | CHIRAMBU | Outpatient surgery activity | Binary |
| FILTRE | CHIMIO | Chemotherapy activity | Binary |
| FILTRE | RTH | Radiotherapy activity | Binary |
| FILTRE | BLOC | Surgery activity | Binary |
| FILTRE | BIO | Medical biology or anatomopathological activity | Binary |
| FILTRE | REA | Intensive care unit | Binary |
| FILTRE | MEDIC | Medication circuit | Binary |
| FILTRE | DOULEUR | Chronic pain | Binary |
| FILTRE | PALIA | Palliative care | Binary |
| FILTRE | CHIRCANCER | Cancer surgery | Binary |
| MCO | SEJHC_MED | Number of inpatient medical stays | Continuous |
| MCO | SEJHC_CHI | Number of inpatient surgery stays | Continuous |
| MCO | SEJHP_MED | Number of outpatient medical stays | Continuous |
| MCO | SEJHP_CHI | Number of outpatient surgery stays | Continuous |
| MCO | LIT_MCO | Number of MCO beds | Continuous |
| BLOCS | SALCHIR | Number of surgery operating rooms | Continuous |
| BLOCS | SALAMBU | Operating rooms dedicated to outpatient surgery | Continuous |
| CANCERO | CANCERO_A1 | Use chemotherapy for cancer treatment | Binary |
| CANCERO | CANCERO_A2 | Use radiotherapy for cancer treatment | Binary |
| CANCERO | CANCERO_A3 | Has an oncology dedicated unit | Binary |
| CANCERO | CANCERO_A11 | Number of patients treated with chemotherapy | Continuous |
| CANCERO | CANCERO_A17 | Number of patients treated with radiotherapy | Continuous |
| - | CANCERO_NB_STAYS_CHIRMED | Number of oncology medical or surgery stays | Continuous |
| - | CANCERO_ACTIVITY | Oncology activity | Continuous |

**Sup. Table 1: List of the variables used for clustering, and their definitions.** All the variables except “*CANCERO_NB_STAYS_CHIRMED*” and “*CANCERO_ACTIVITY*” are coming from SAE. The variables are either binary or continuous. Oncology activity is the sum of “CANCERO_NB_STAYS_CHIRMED”, “*CANCERO_A17*” and “*CANCERO_A11”.*

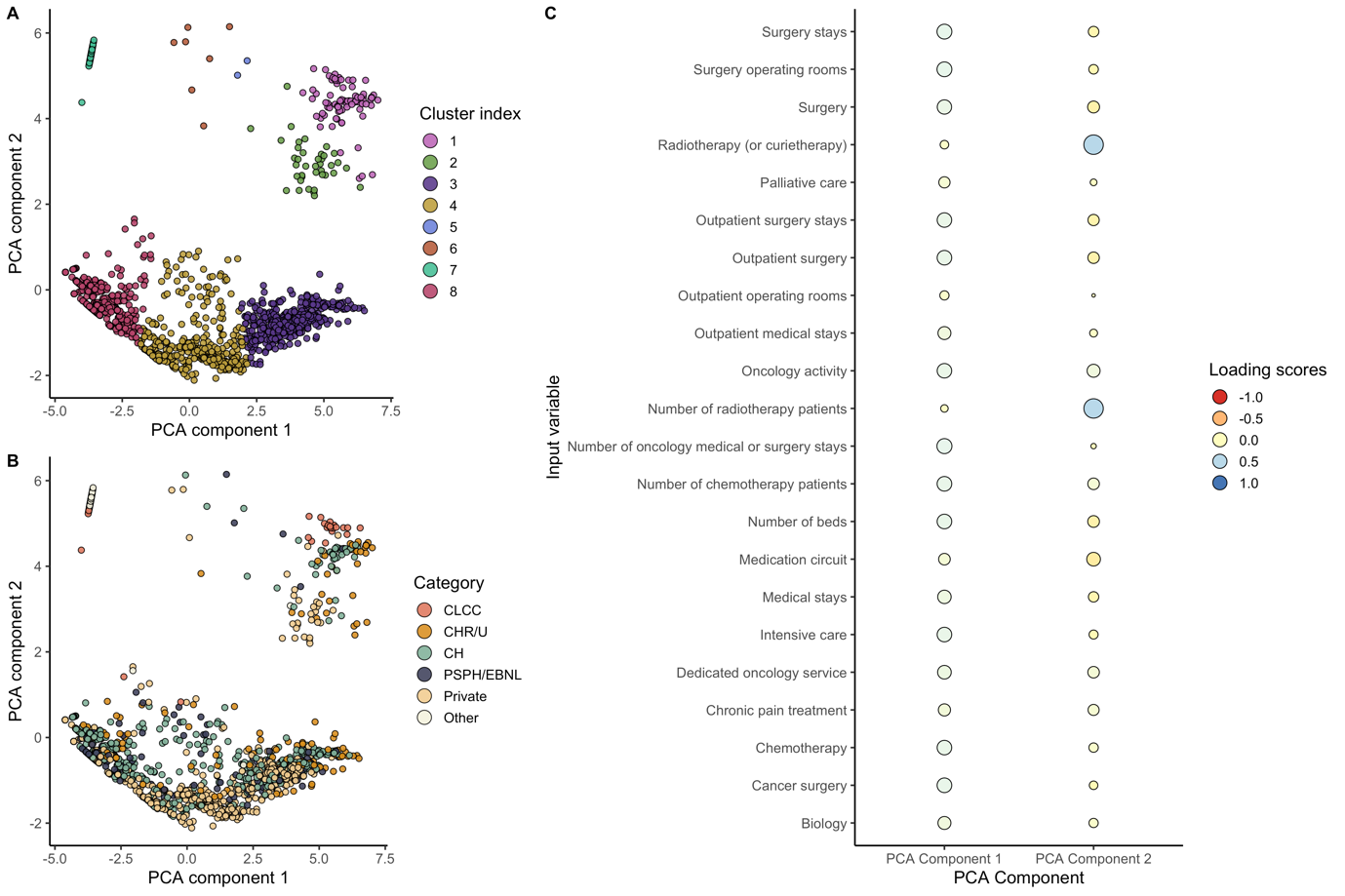

**Sup. Figure 1: Principal Component Analysis (PCA) interpretation.** Care centers are showed as points in the 2-dimensional PCA space. Points are colored by cluster index (A) and hospital type (B). CLCC care centers are close together in the PCA space, proving they have similar activity and services distribution. PCA components are a linear combination of the input variables (C). The loading scores reflect how much the input variable contributed to the PCA component. Component 1 is associated with most of the variables, while component 2 is linked with radiotherapy variables. Hence, we interpret component 1 as hospital size and component 2 as oncology specialization.

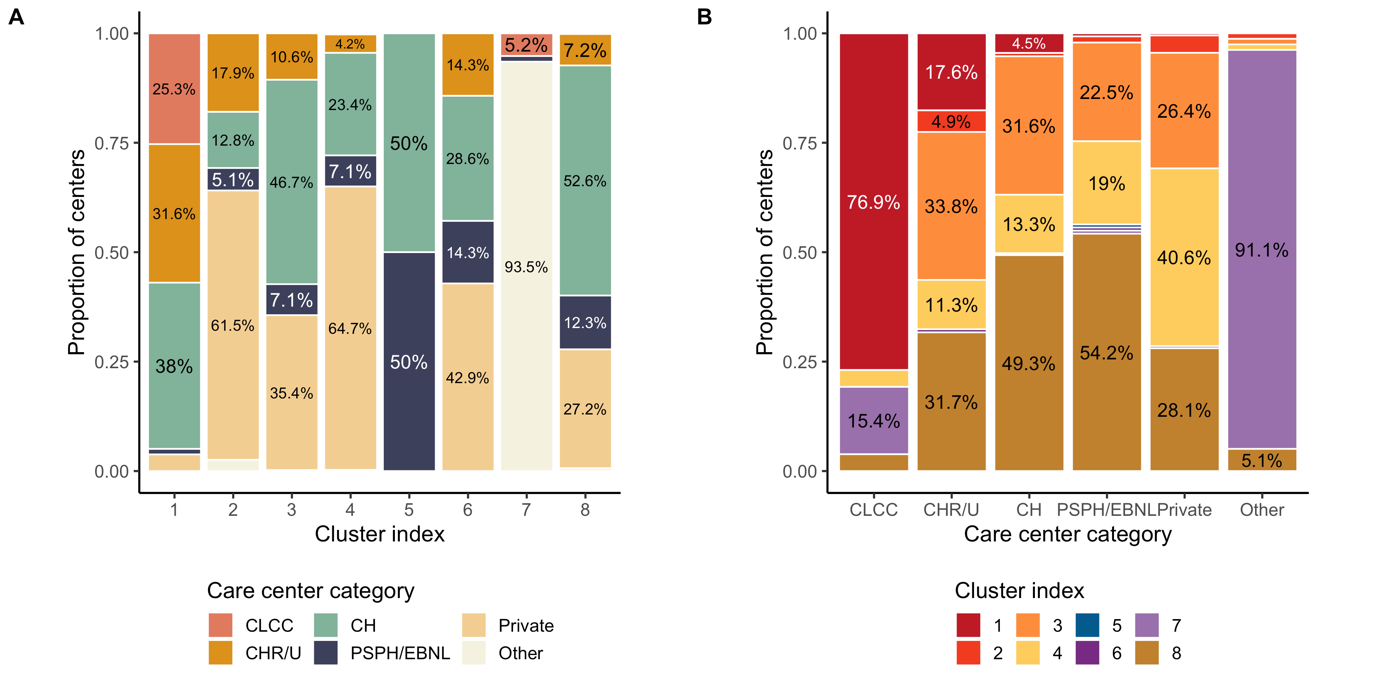

**Sup. Figure 2: Comparison between hospital types and assigned clusters.** The majority of the CLCC care centers are grouped together in cluster 1. Moreover, cluster 1 has a very low percentage of private hospitals, whereas this proportion is the much higher in cluster 2. “Other” care centers are mostly private practice radiotherapy structures, and they are regrouped in cluster 7.

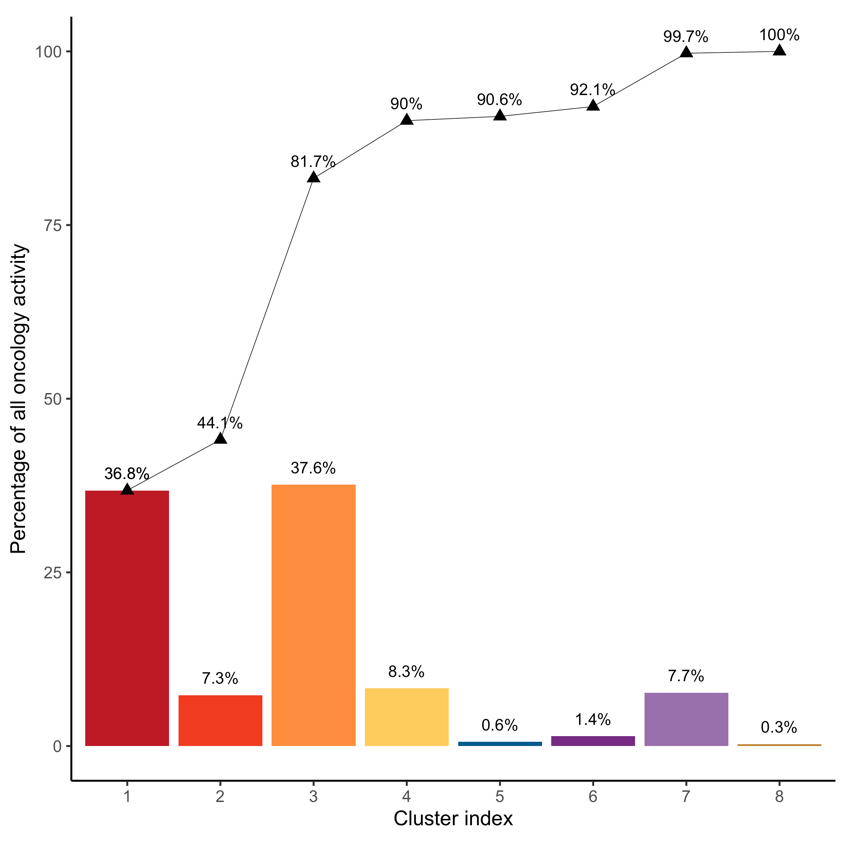

**Sup. Figure 3: Cumulative sum of the oncology activity, per cluster.** Most of the oncology activity is handled by care centers from clusters 1 and 3. While there are only n=79 care centers in cluster 1, their total activity is almost as large as the n=451 care centers from cluster 3.

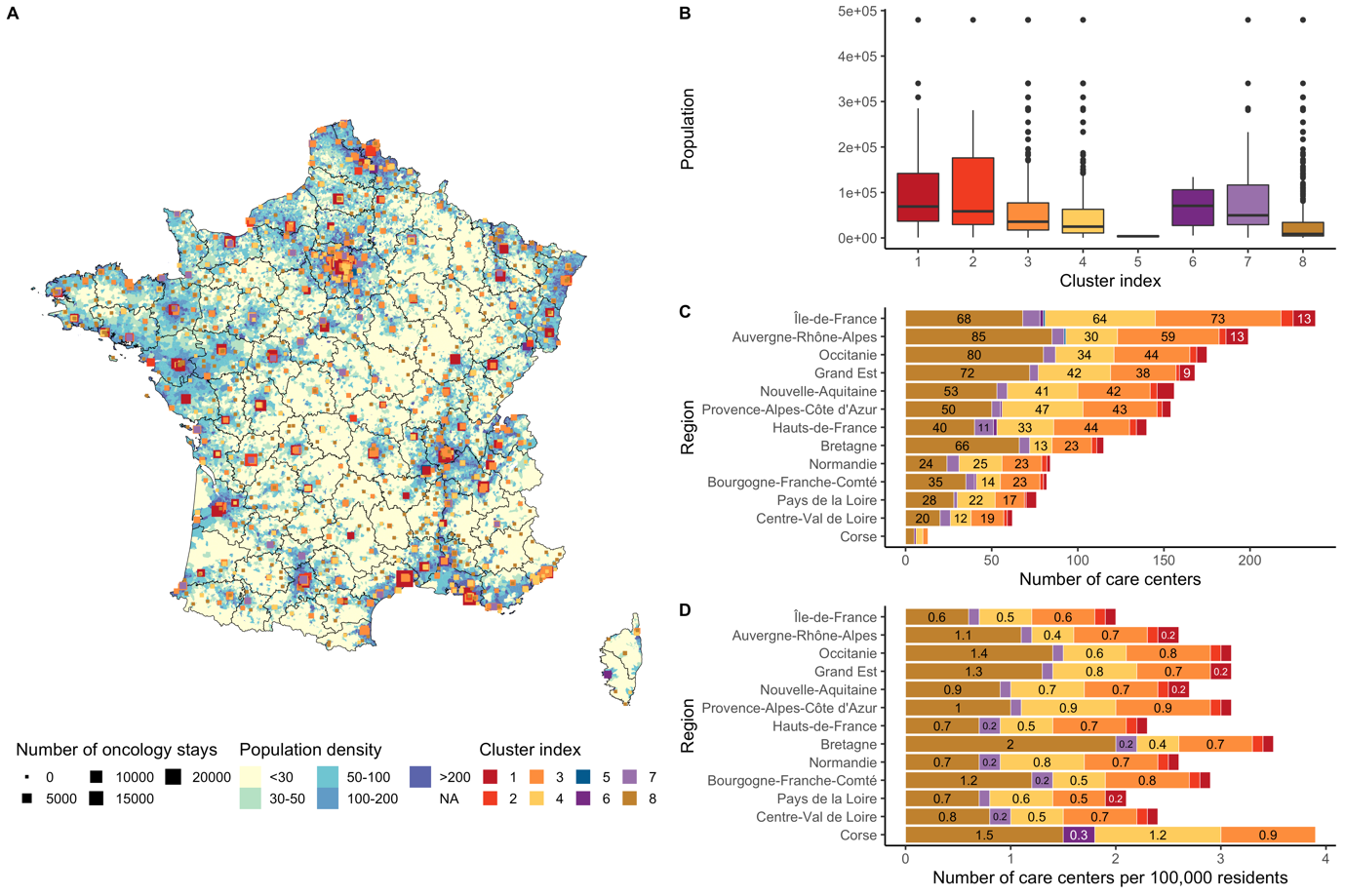

**Sup. Figure 4: Care centers spatial distribution, compared with population density.** Population density in metropolitan France is unevenly distributed across the country (A). Areas in the middle, near the Pyrenees and the Alps have very low population densities. The most specialized care centers are in dense areas and in large municipalities (B). While Ile-de-France has the highest number of care centers, it has the least care centers per 100,000 habitants.

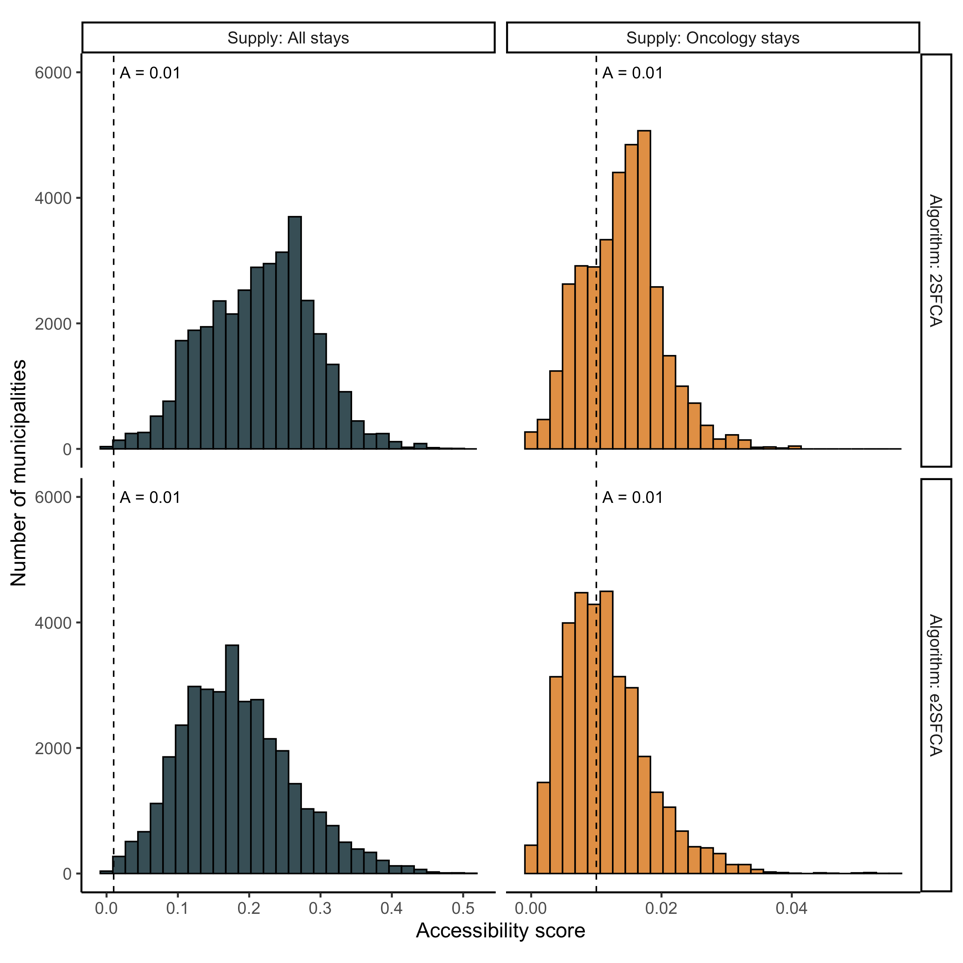

**Sup. Figure 5: Comparison of accessibility distributions, for different algorithms and supply variables.** We compared accessibility distributions with 2SFCA and e2SFCA algorithms. We also tried two variables as supply: all medical surgery or obstetric (MCO) stays, versus oncology activity. We notice that accessibility is lower when using oncology activity as supply rather than MCO stays. We also see that accessibility from e2SFCA algorithm is lower than 2SFCA, due to the distance decay.

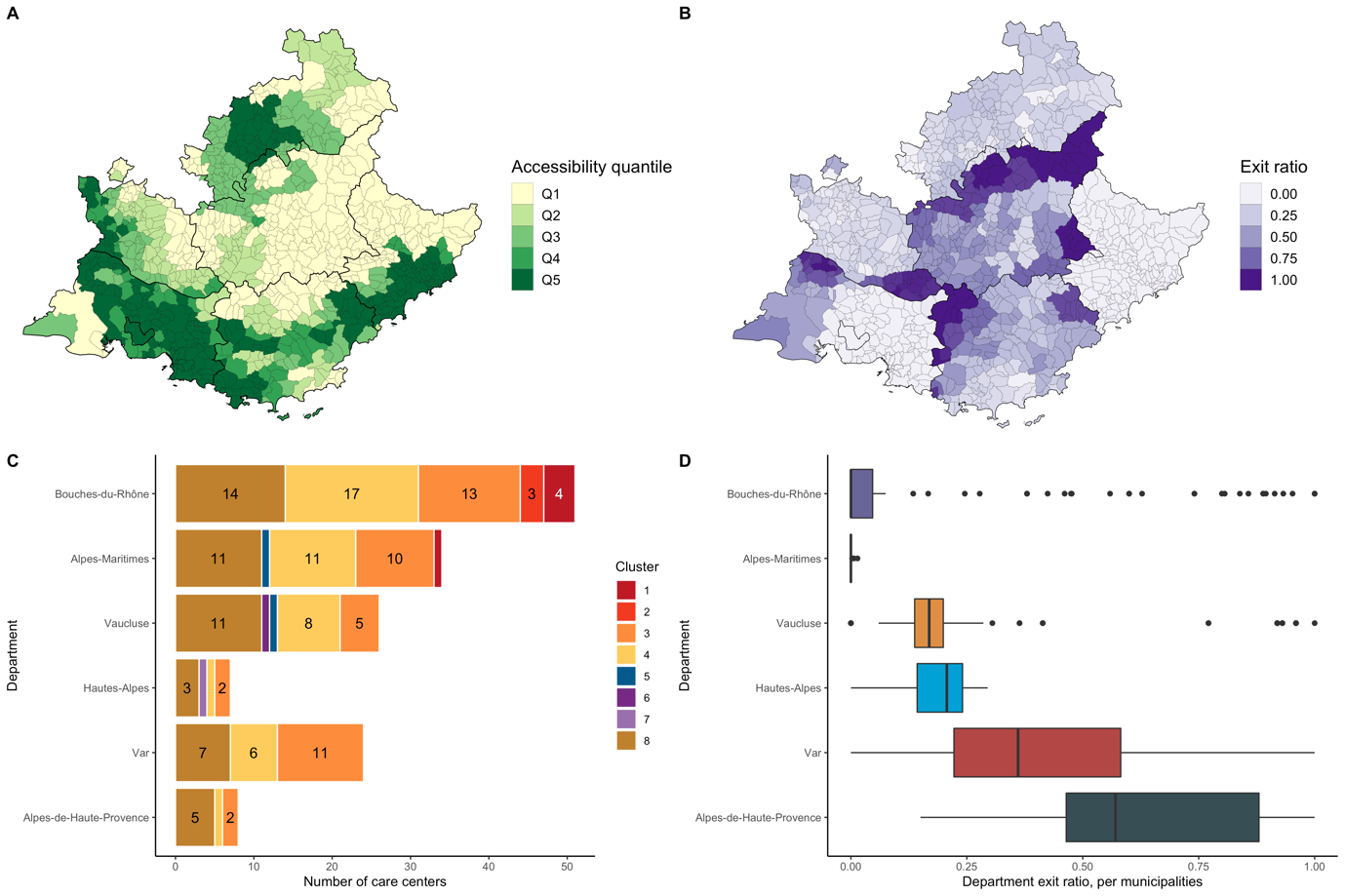

**Sup. Figure 6: Comparison between accessibility score and department exit ratio by municipality.** Department exit ratio is defined as the percentage of patients from a municipality who visited a care center from another department. Exit ratio is higher near departments borders. Moreover, exit ratio is higher in municipalities with low accessibility scores (A, B). For instance, exit ratio distribution in Alpes-de-Haute-Provence is much higher than Bouches-du-Rhone (D). Exit ratio is very low in Alpes-Maritimes, even though many of its municipalities have poor accessibility: exit ratio is heavily influenced by the presence of specialized care centers in the department.

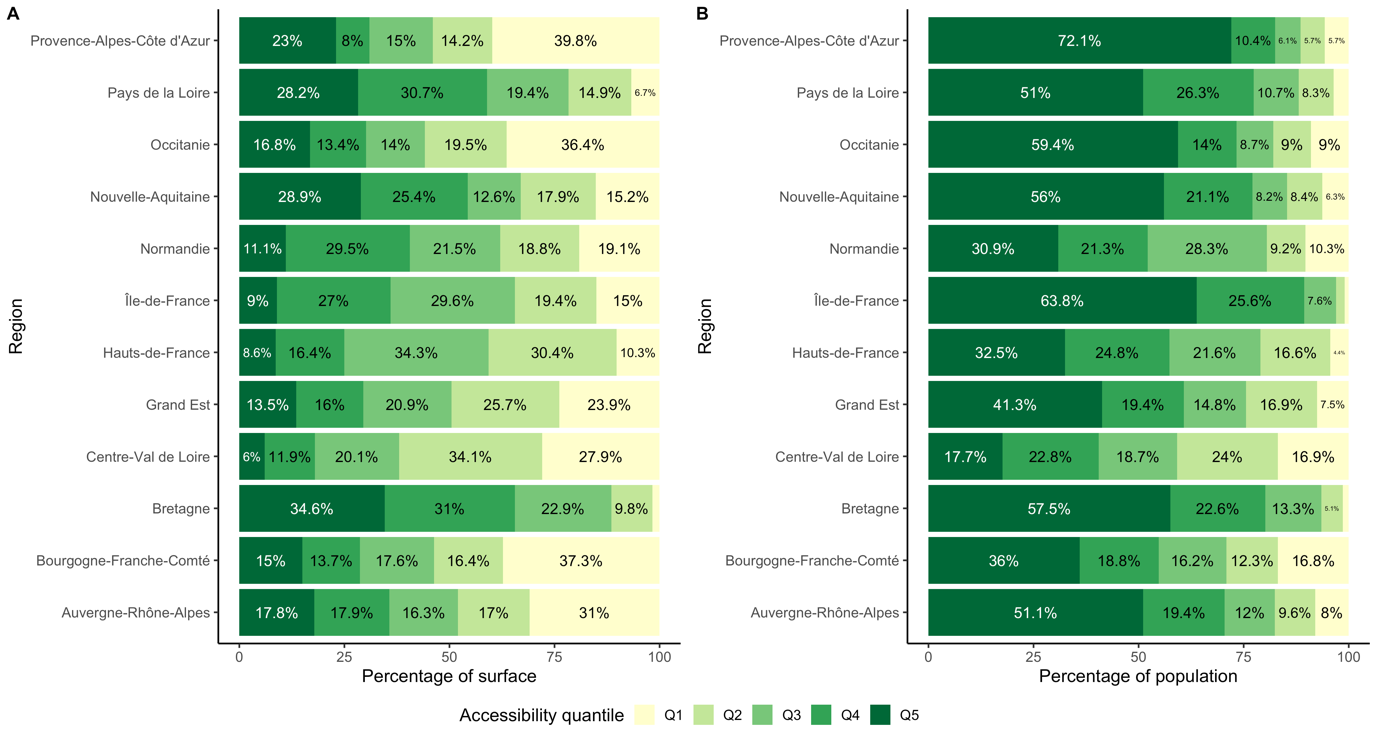

**Sup. Figure 7: Percentage of surface and population by accessibility quantile.** We compared the accessibility quantiles distribution per region. Plot (A) shows the percentage of the regions’ surface per quantile, while plot (B) shows the percentage of population per quantile. In Ile-de-France, 15% of the surface belongs to Q1 accessibility, yet the corresponding population is very low. The percentage of population living in Q1 quantile is the highest in Centre-Val-de-Loire (16.9%) and Bourgogne-Franche-Comté (16.8%).

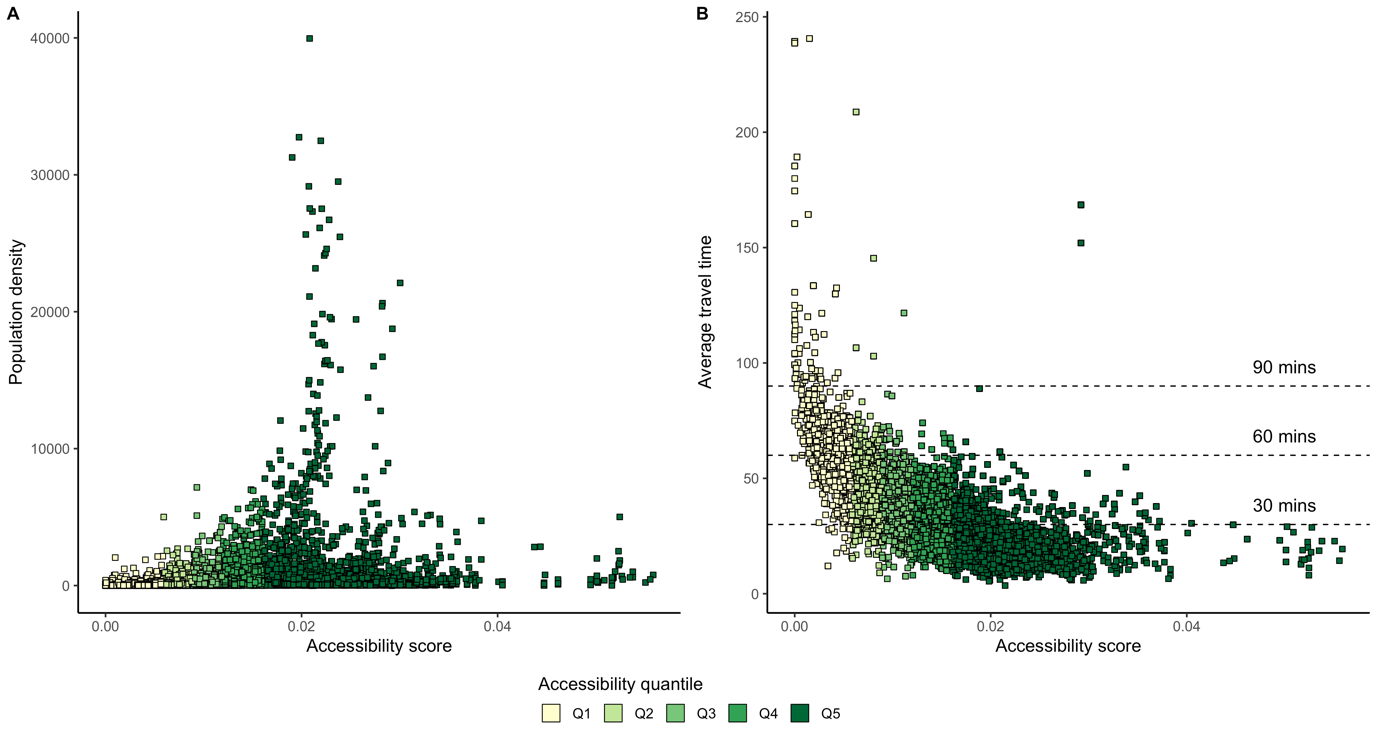

**Sup. Figure 8: Accessibility compared with population density and average travel time.** The dense municipalities in metropolitan France have a median accessibility around 0.02 (A). Very low or very high accessibility scores are attributed to low-density municipalities. The average travel time for patients living in Q5 accessibility areas is lower than 30 minutes most of the time. For patients living in Q1 areas, it’s unlikely that the average travel time falls under 1 hour.

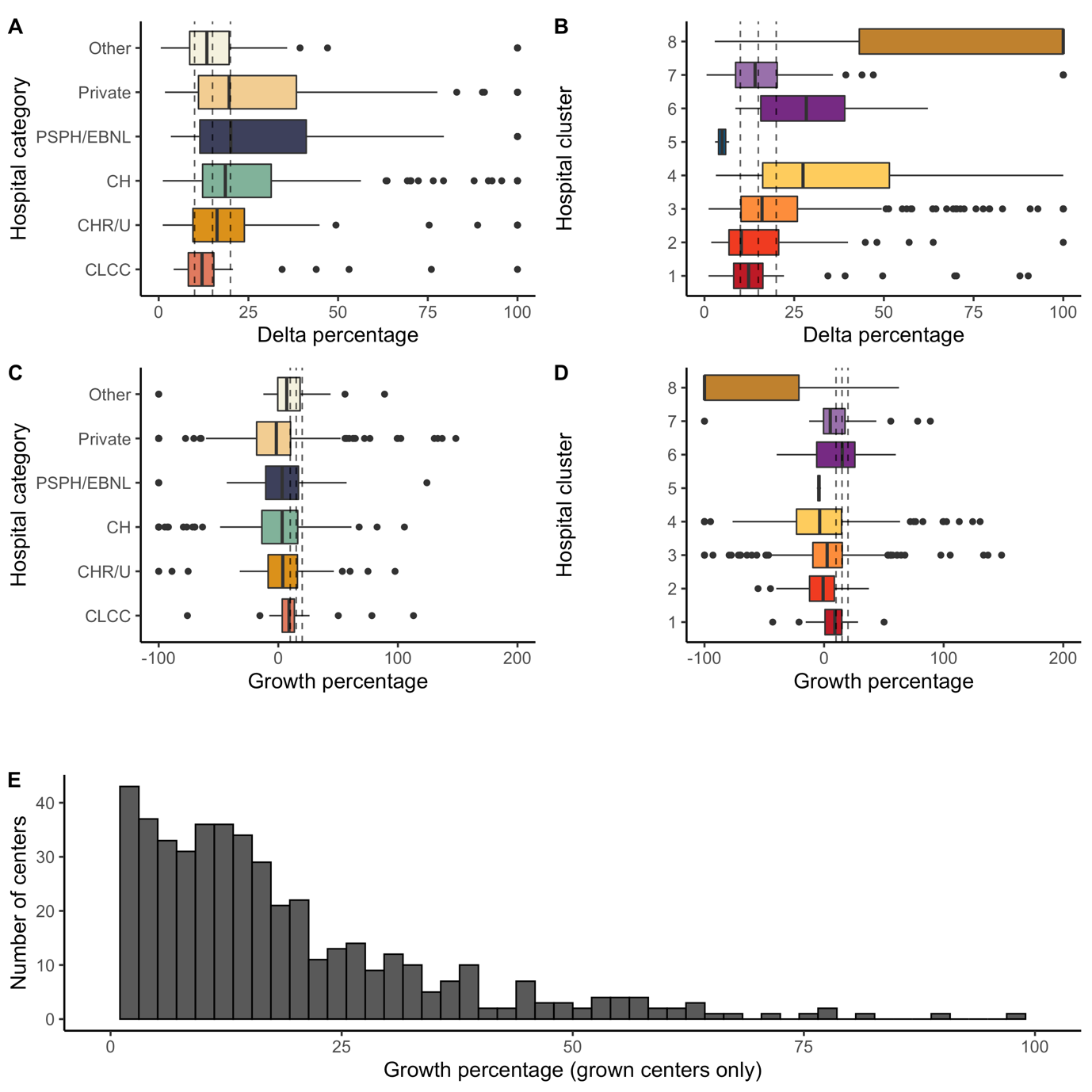

**Sup. Figure 9: Oncology activity evolution, between 2016 and 2019.** Plots (A) and (B) show the activity delta percentage: $\frac{Activity_{max}-Activity_{min}}{Activity_{max}}$; plots (C) and (D) show the growth percentage: $\frac{Activity_{2019}-Activity_{2016}}{Activity_{2016}}$. The delta percentage is an indicator for capacity variation, while growth percentage shows activity growth. If the growth percentage is -100%, the care center was emptied between 2016 and 2019. Three dashed lines are drawn on each plot, corresponding to 10%, 15% and 20%. Plot (E) shows the distribution of the care center growth percentage, for hospitals that grew and with initial oncology activity. The mean growth percentage is 23% and was used as maximum growth percentage in the optimization algorithm.

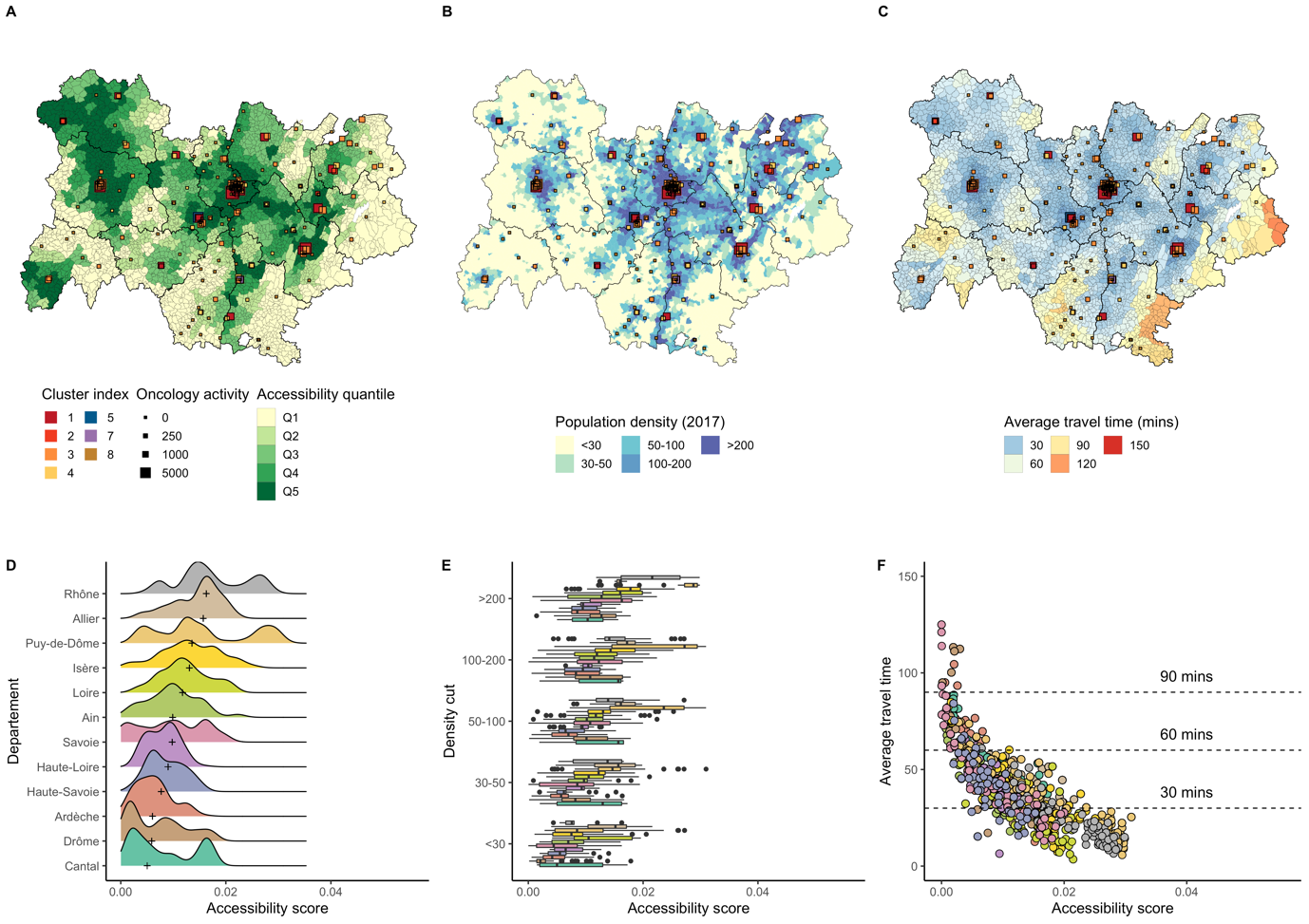

**Sup. Figure 10: Accessibility distribution in Auvergne-Rhone-Alpes.**

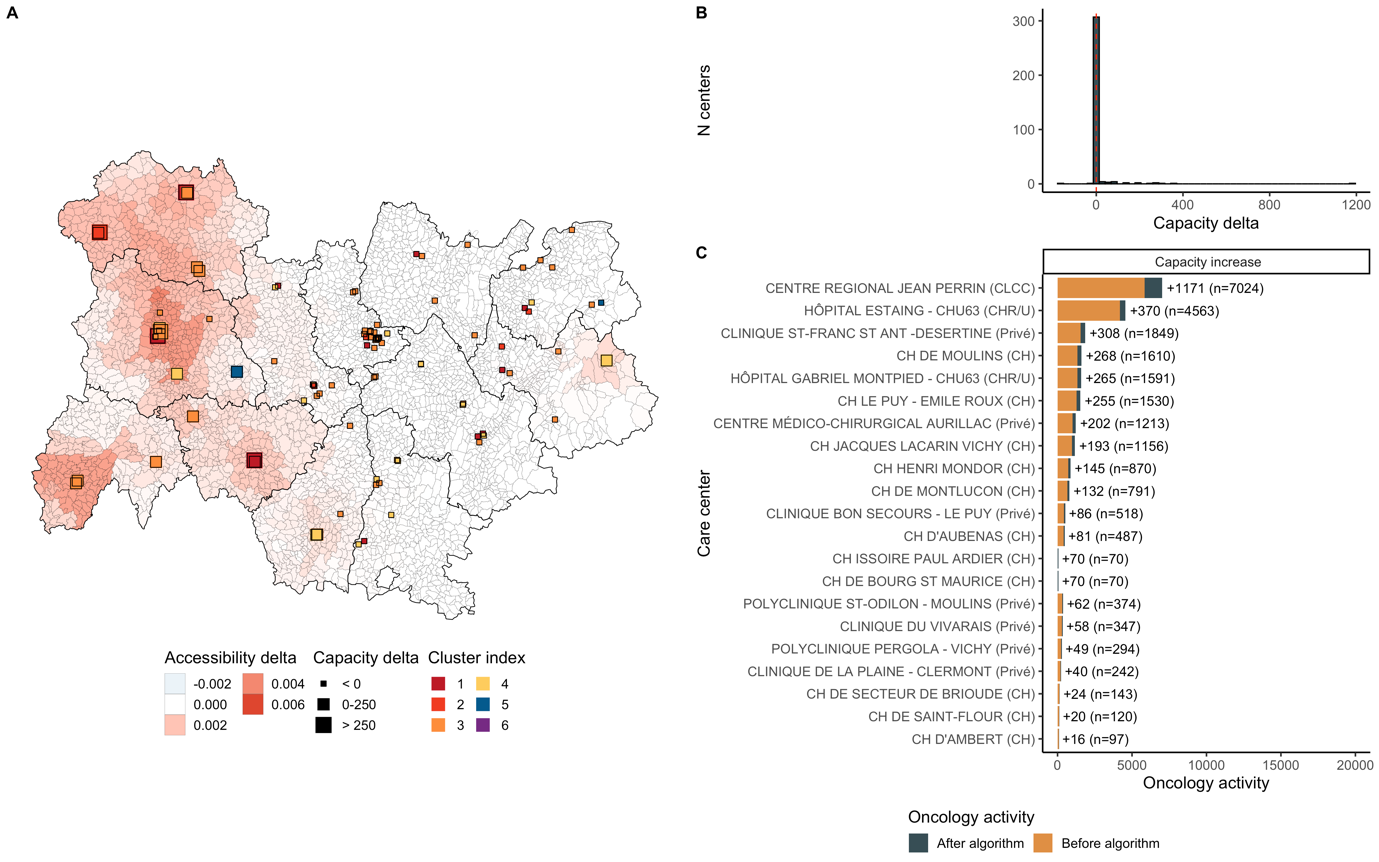

**Sup. Figure 11: Optimization results in Auvergne-Rhone-Alpes.** Additional activity was 3,883. 23 centers grew and 2 decreased. Median accessibility before optimization was 0.0092 and 0.0095 after, corresponding to a 3.2% increase. Accessibility grew around Moulins, Montluçon, Le Puy en Velay, Clermont-Ferrand and Aurillac.

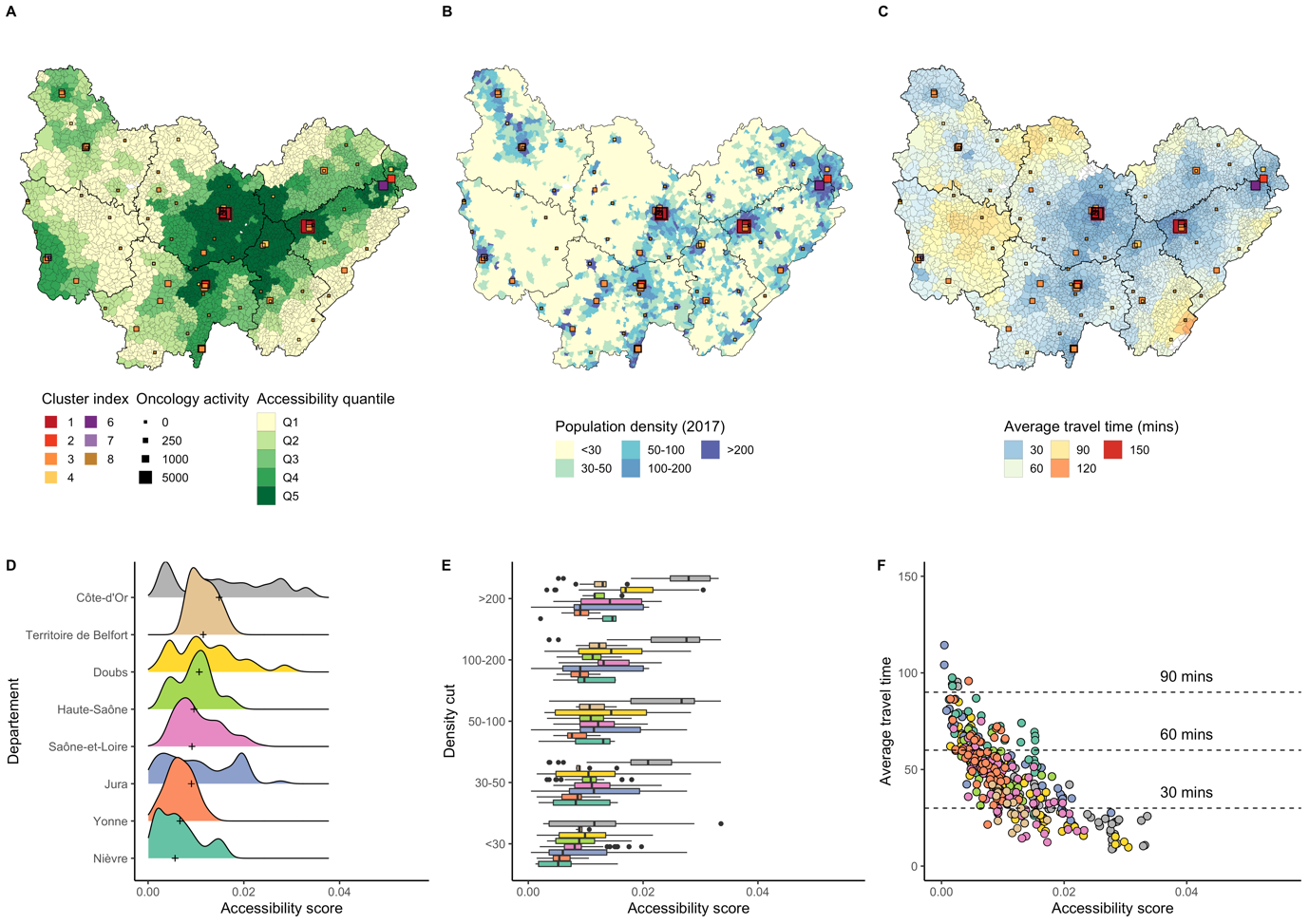

**Sup. Figure 12: Accessibility distribution in Bourgogne-Franche-Comté.**

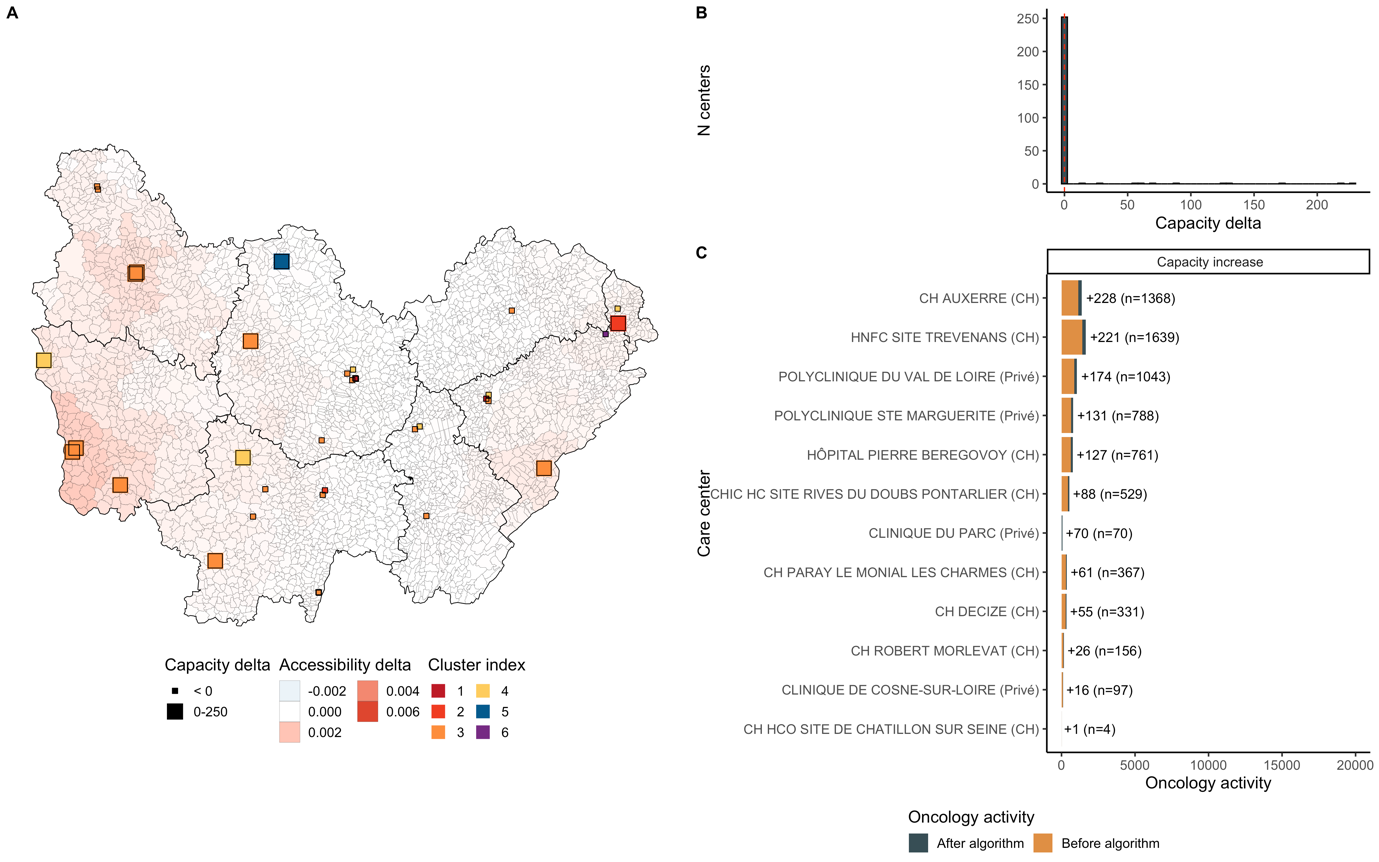

**Sup. Figure 13: Optimization results in Bourgogne-Franche-Comté.** Additional activity was 1,330. 13 centers grew and 0 decreased. Median accessibility before optimization was 0.0096 and 0.0098 after, corresponding to a 1.9% increase. Accessibility grew around Nevers, Belfort, Vesoul and Auxerre.

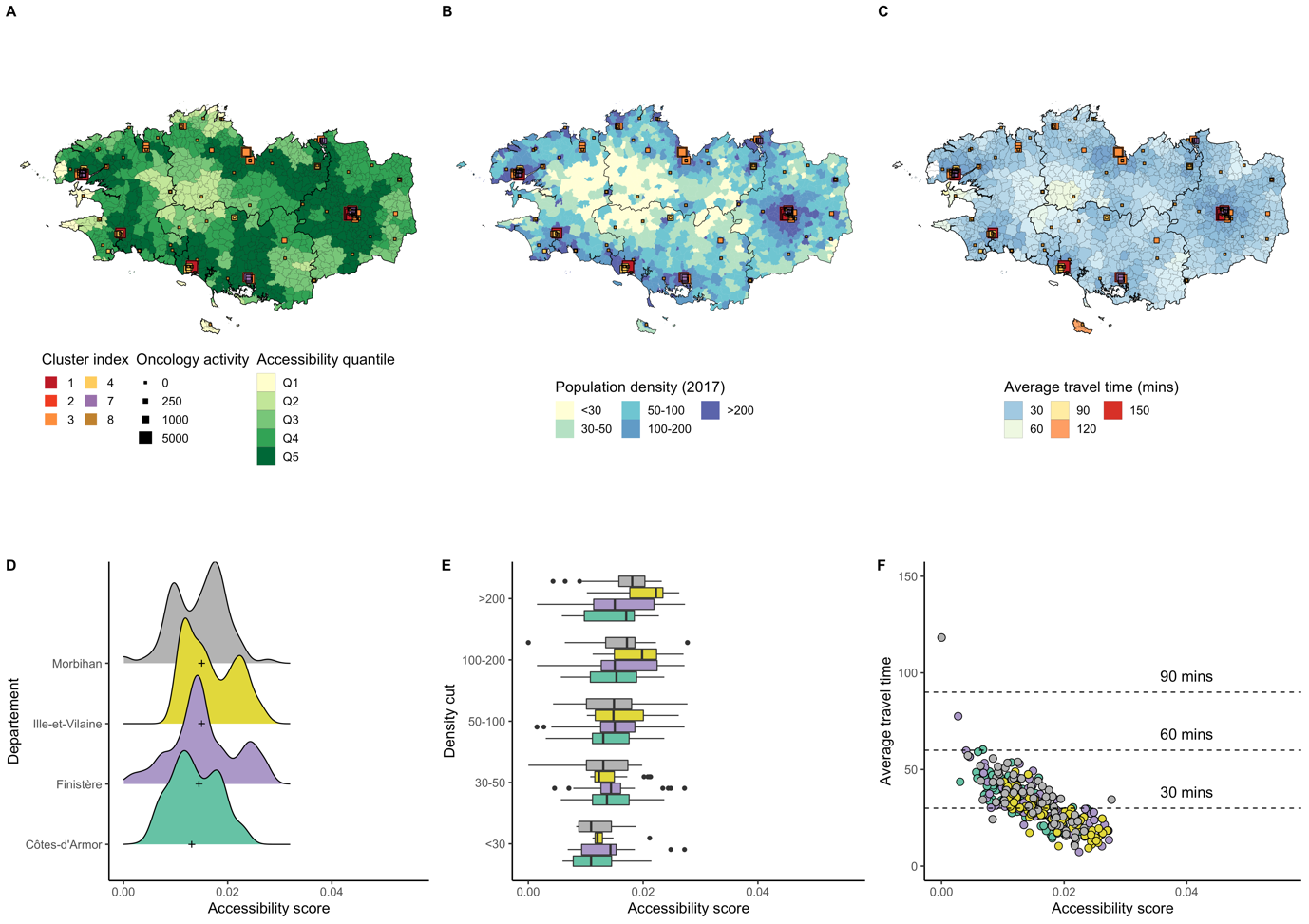

**Sup. Figure 14: Accessibility distribution in Bretagne.**

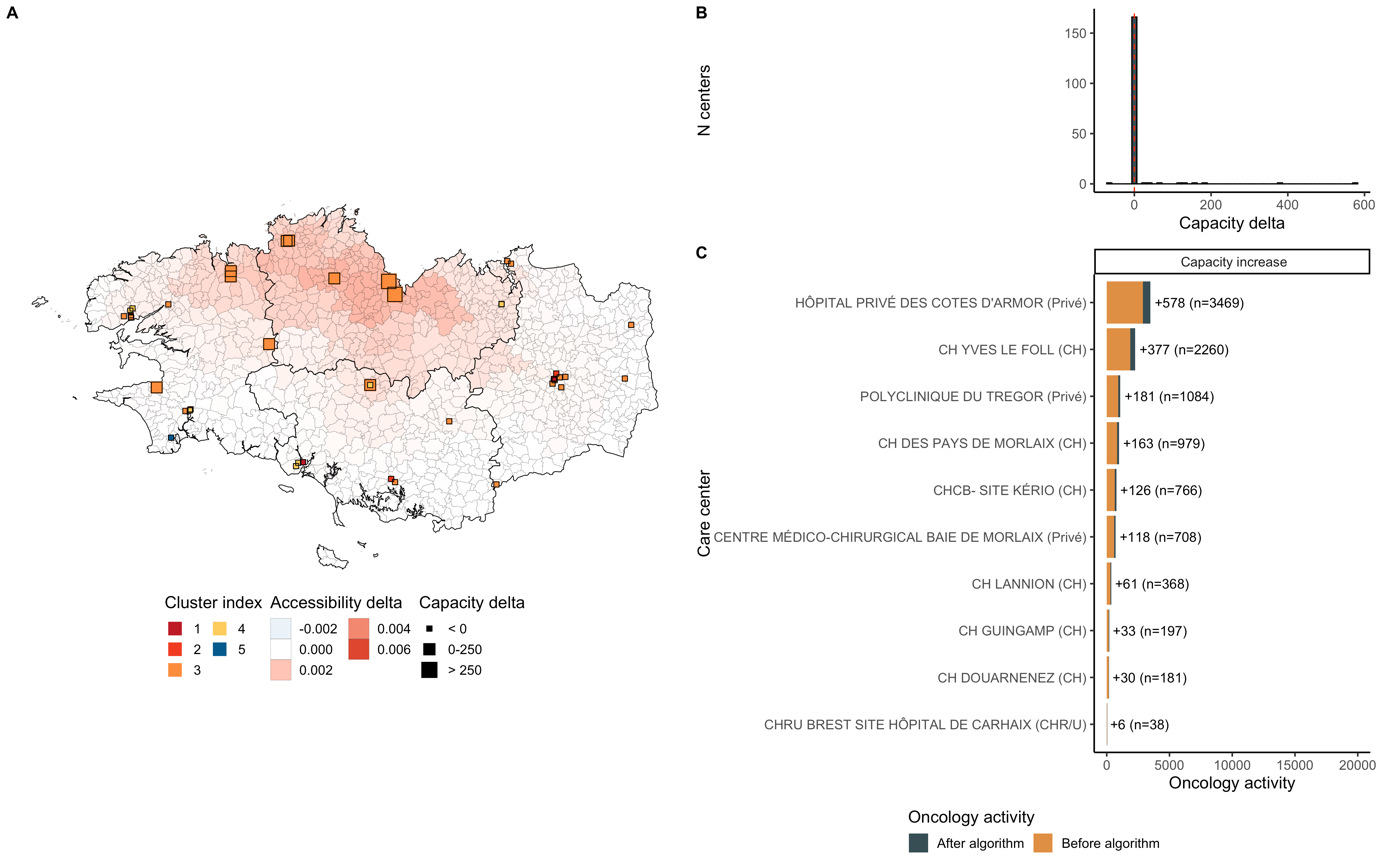

**Sup. Figure 15: Optimization results in Bretagne.** Additional activity was 1,773. 10 centers grew and 2 decreased. Median accessibility before optimization was 0.0131 and 0.0134 after, corresponding to a 2.4% increase. Accessibility grew around St-Brieuc and Quimper.

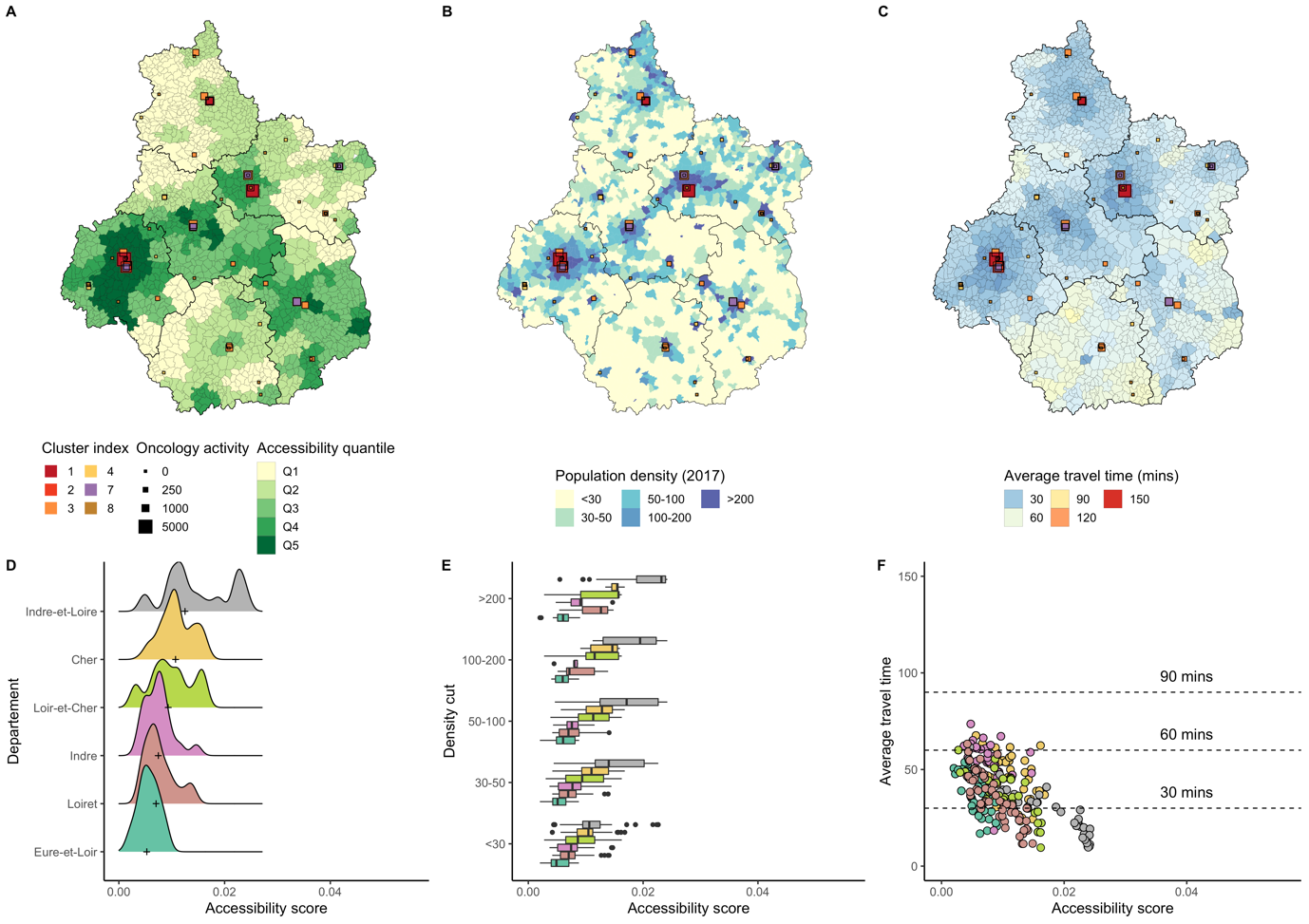

**Sup. Figure 16: Accessibility distribution in Centre-Val-de-Loire.**

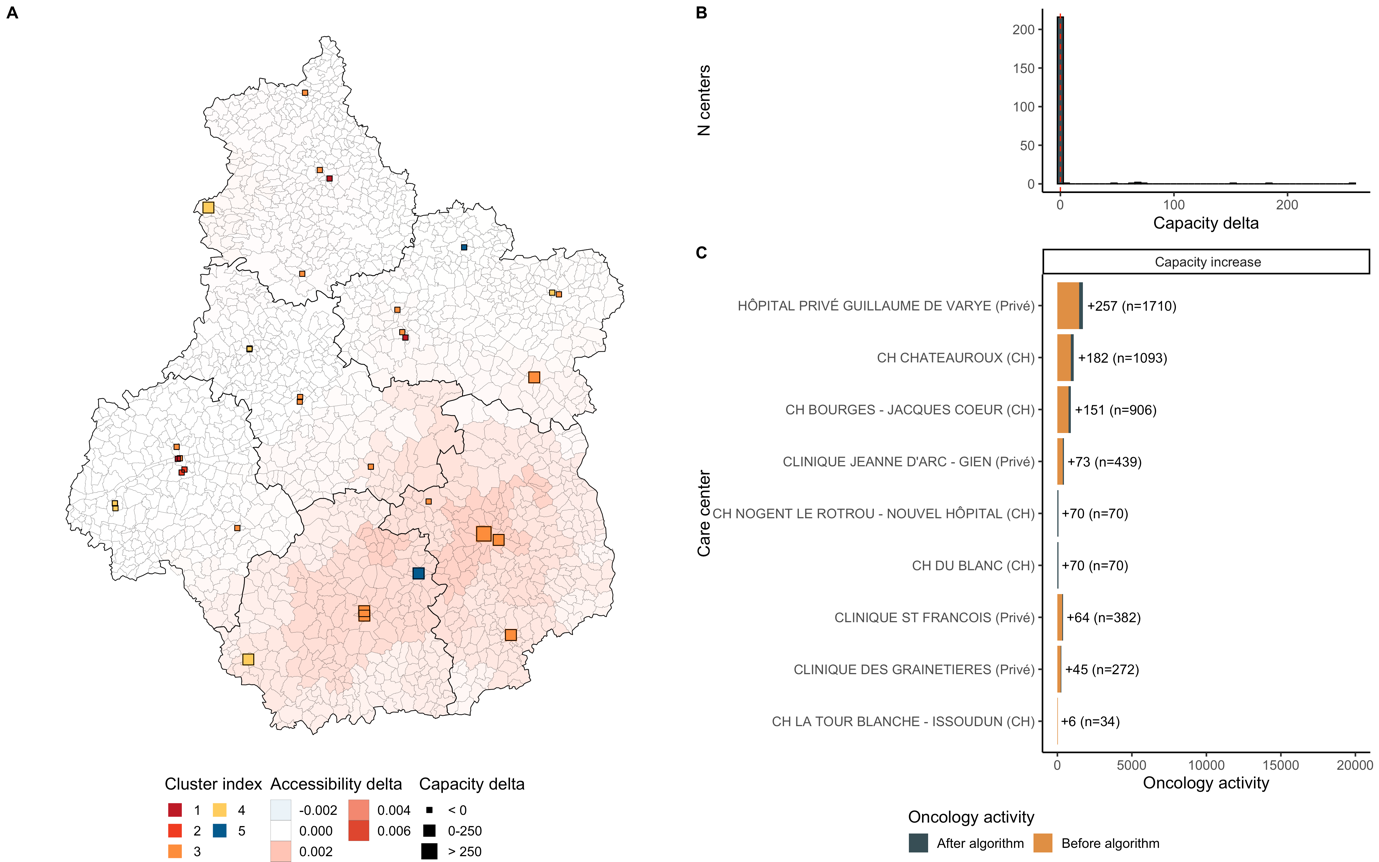

**Sup. Figure 17: Optimization results in Centre-Val-de-Loire.** Additional activity was 1,072. 10 centers grew and 1 decreased. Median accessibility before optimization was 0.0099 and 0.0102 after, corresponding to a 2.9% increase. Accessibility grew around Tours, Blois, Bourges and Chateauroux.

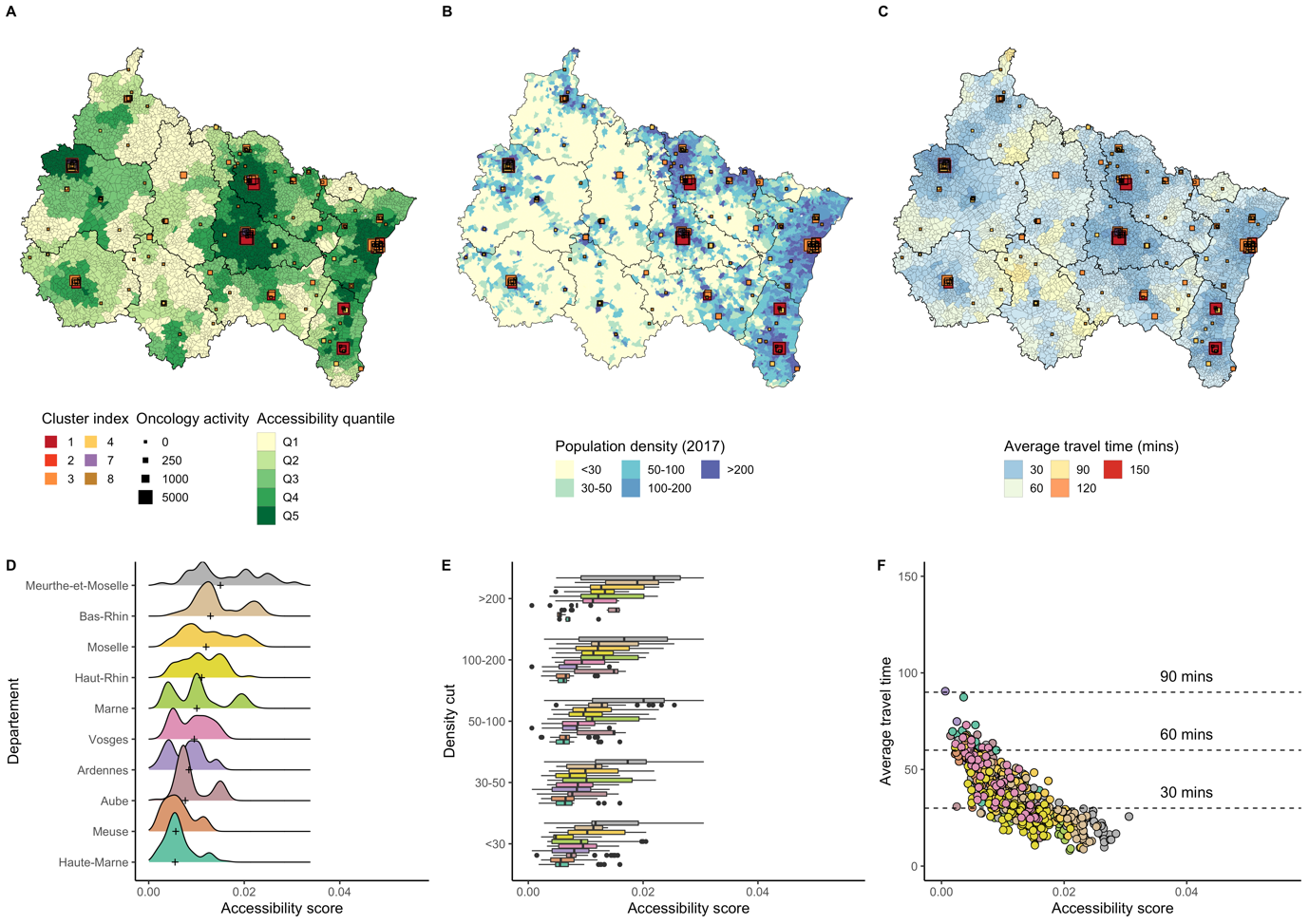

**Sup. Figure 18: Accessibility distribution in Grand-Est.**

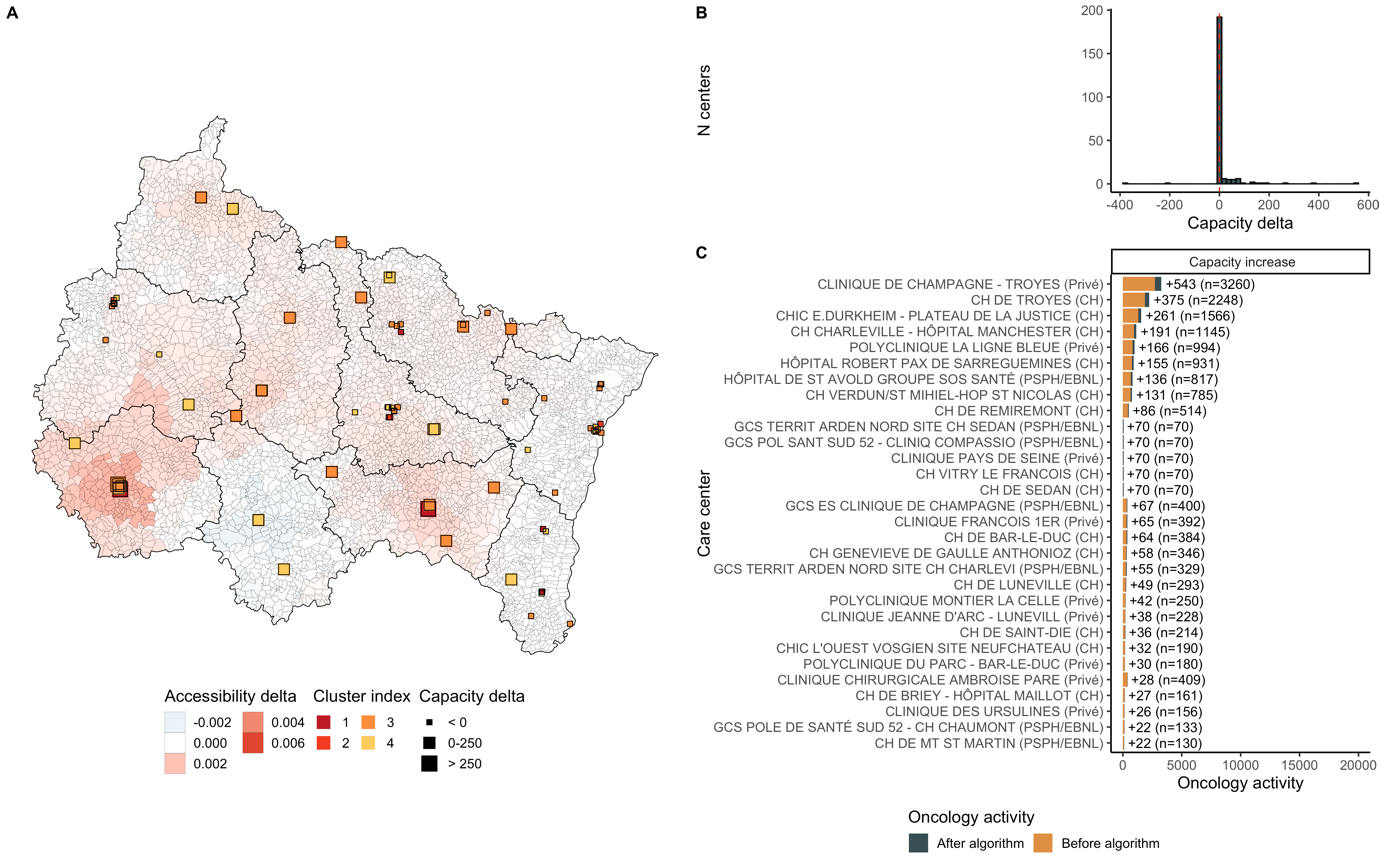

**Sup. Figure 19: Optimization results in Grand-Est.** Additional activity was 2,663. 31 centers grew and 4 decreased. Median accessibility before optimization was 0.0096 and 0.0099 after, corresponding to a 3% increase. Accessibility grew around Troyes and Epinal.

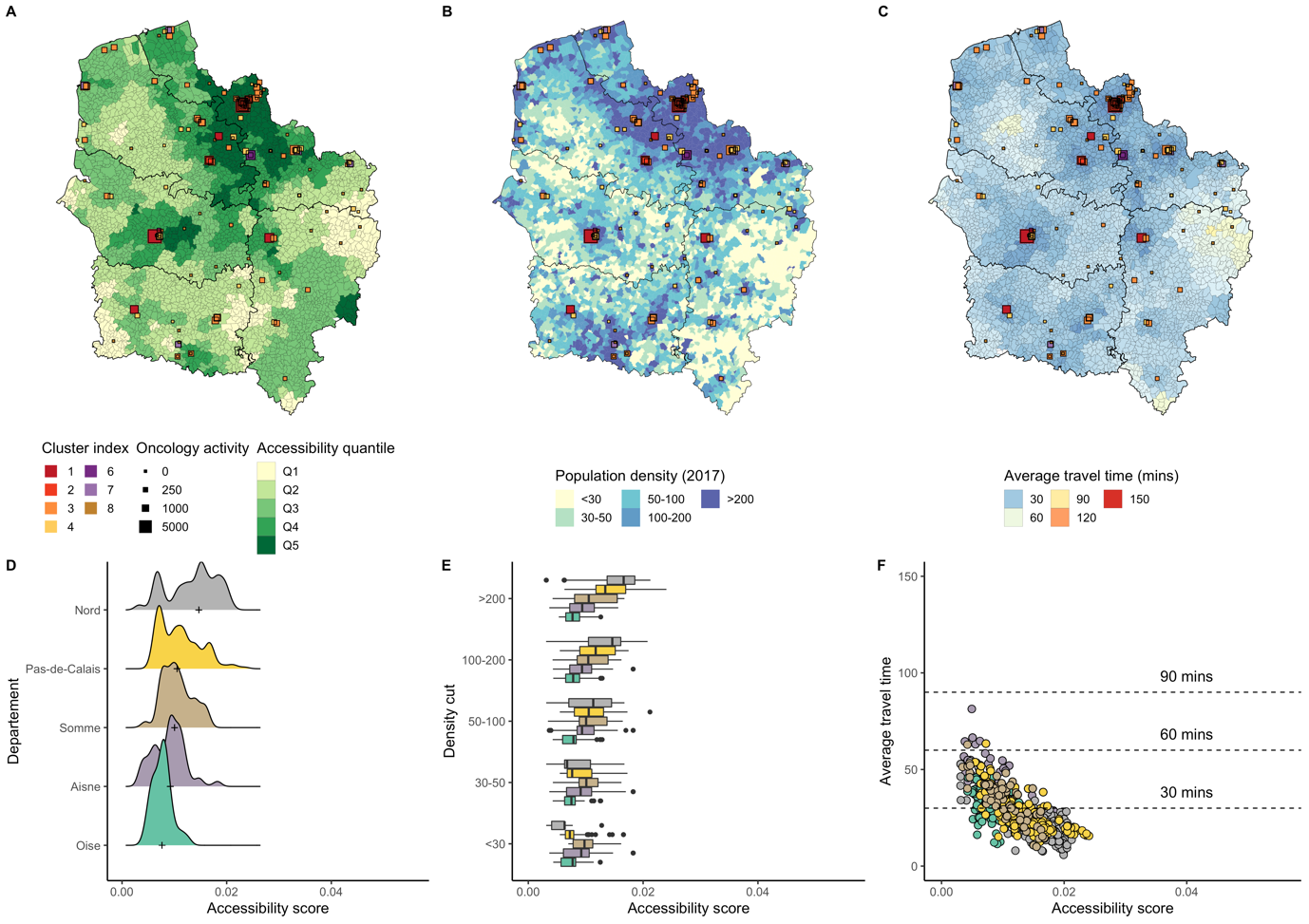

**Sup. Figure 20: Accessibility distribution in Hauts-de-France.**

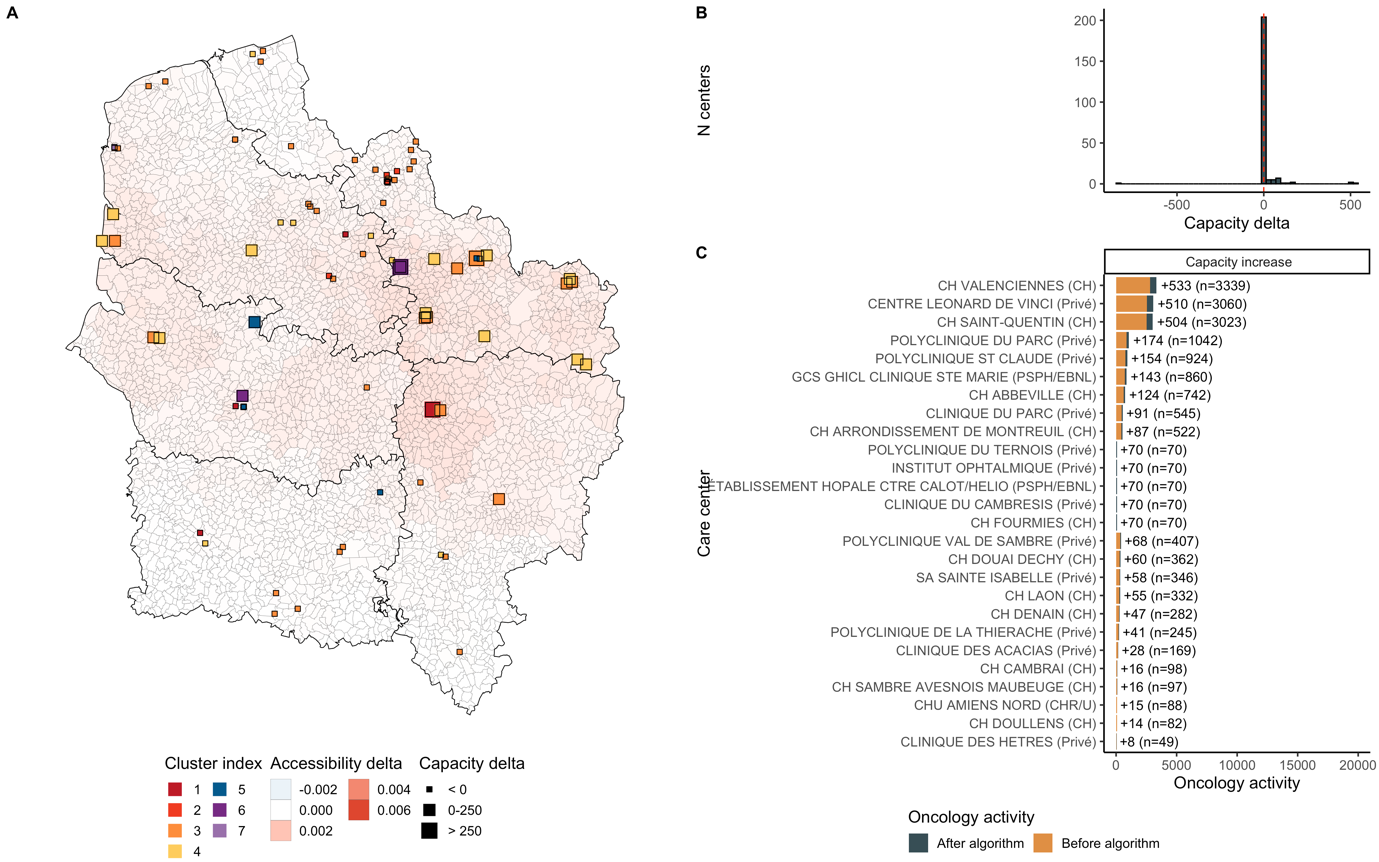

**Sup. Figure 21: Optimization results in Hauts-de-France.** Additional activity was 2,520. 29 centers grew and 1 decreased. Median accessibility before optimization was 0.01 and 0.0102 after, corresponding to a 2.1% increase. Accessibility grew around St-Quentin and Valenciennes.

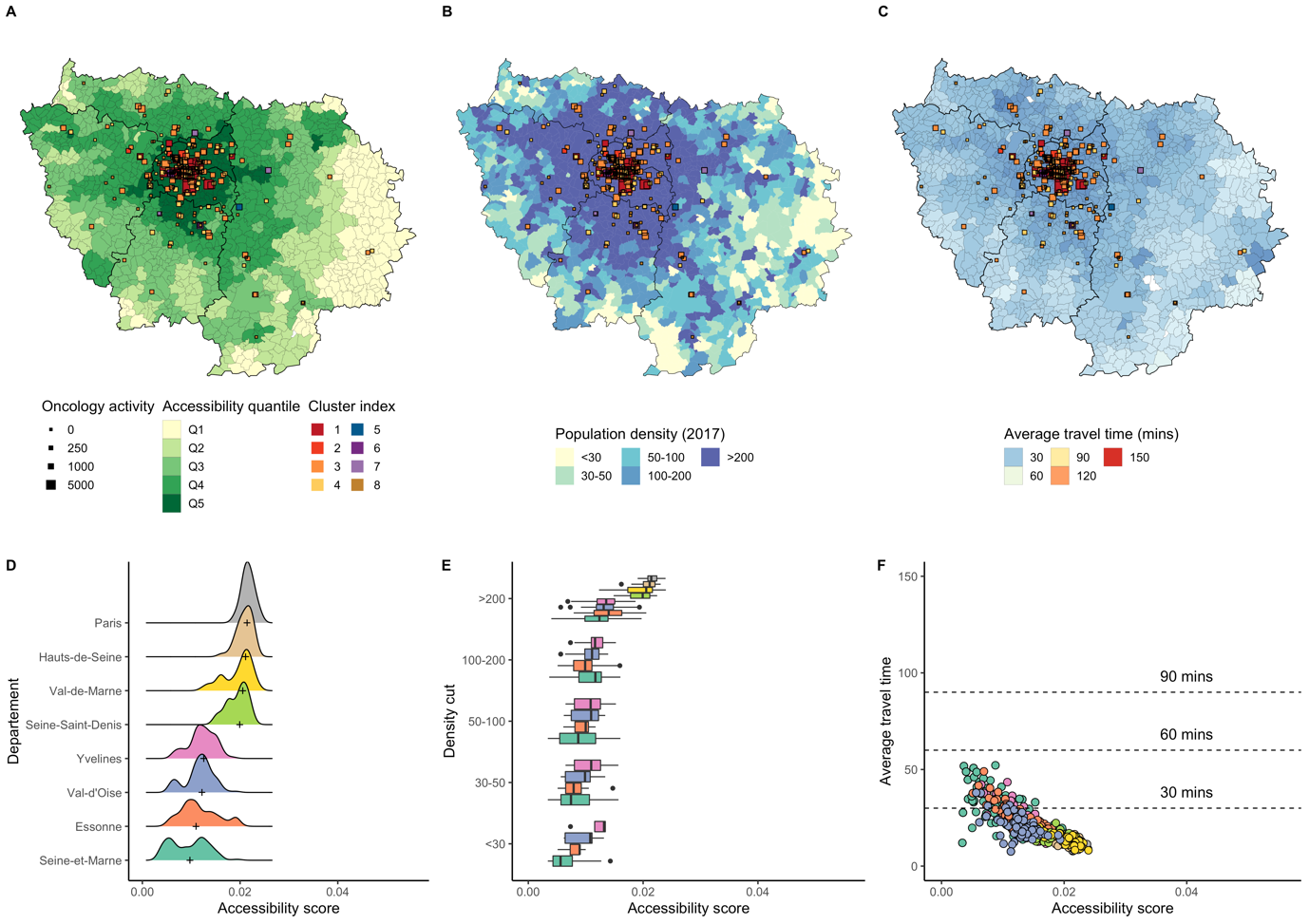

**Sup. Figure 22: Accessibility distribution in Ile-de-France.**

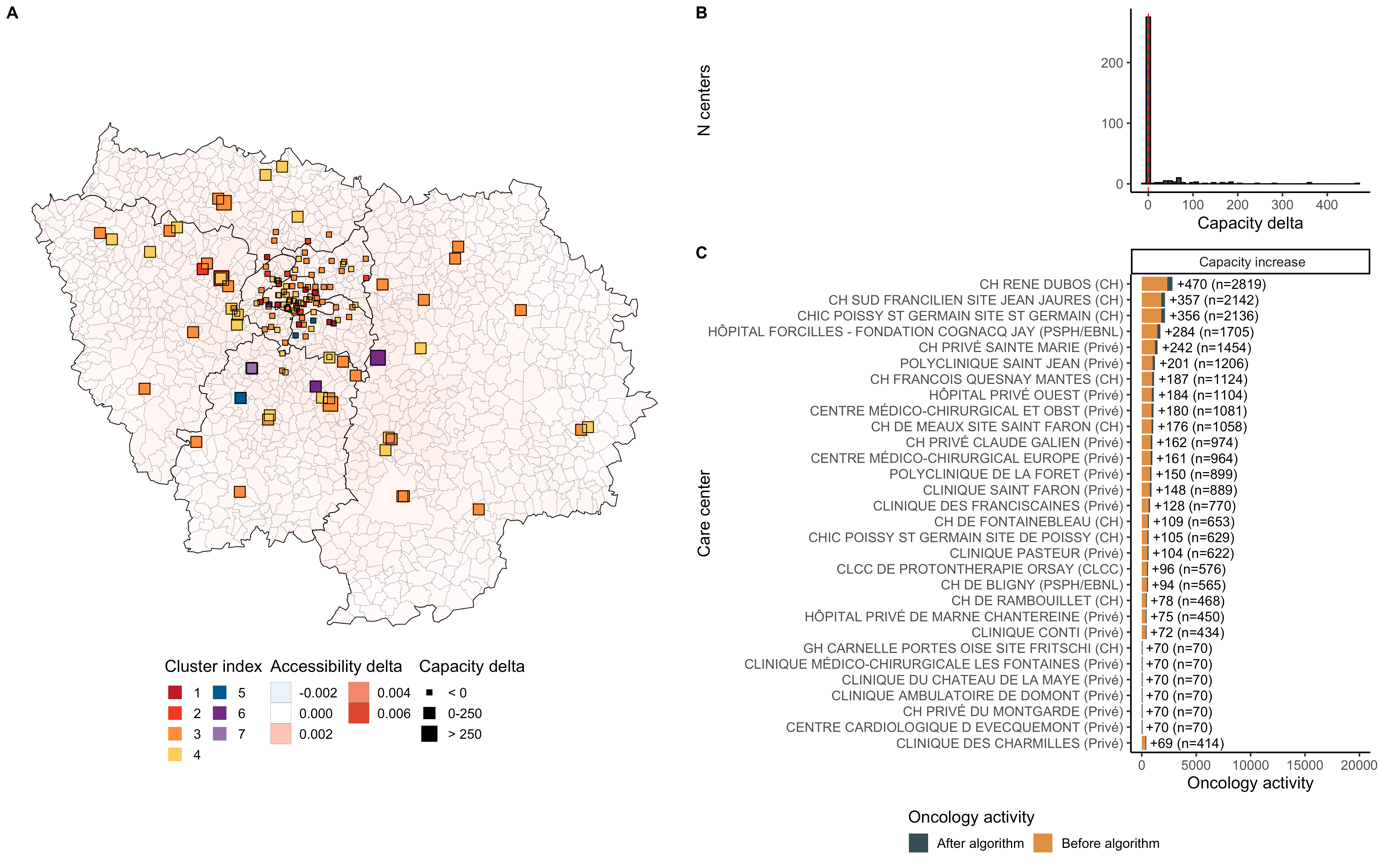

**Sup. Figure 23: Optimization results in Ile-de-France.** Additional activity was 5,826. 44 centers grew and 1 decreased. Median accessibility before optimization was 0.0088 and 0.0089 after, corresponding to a 1.3% increase. Accessibility grew around Mantes-la-Jolie, Rambouillet, Melun, and Evry.

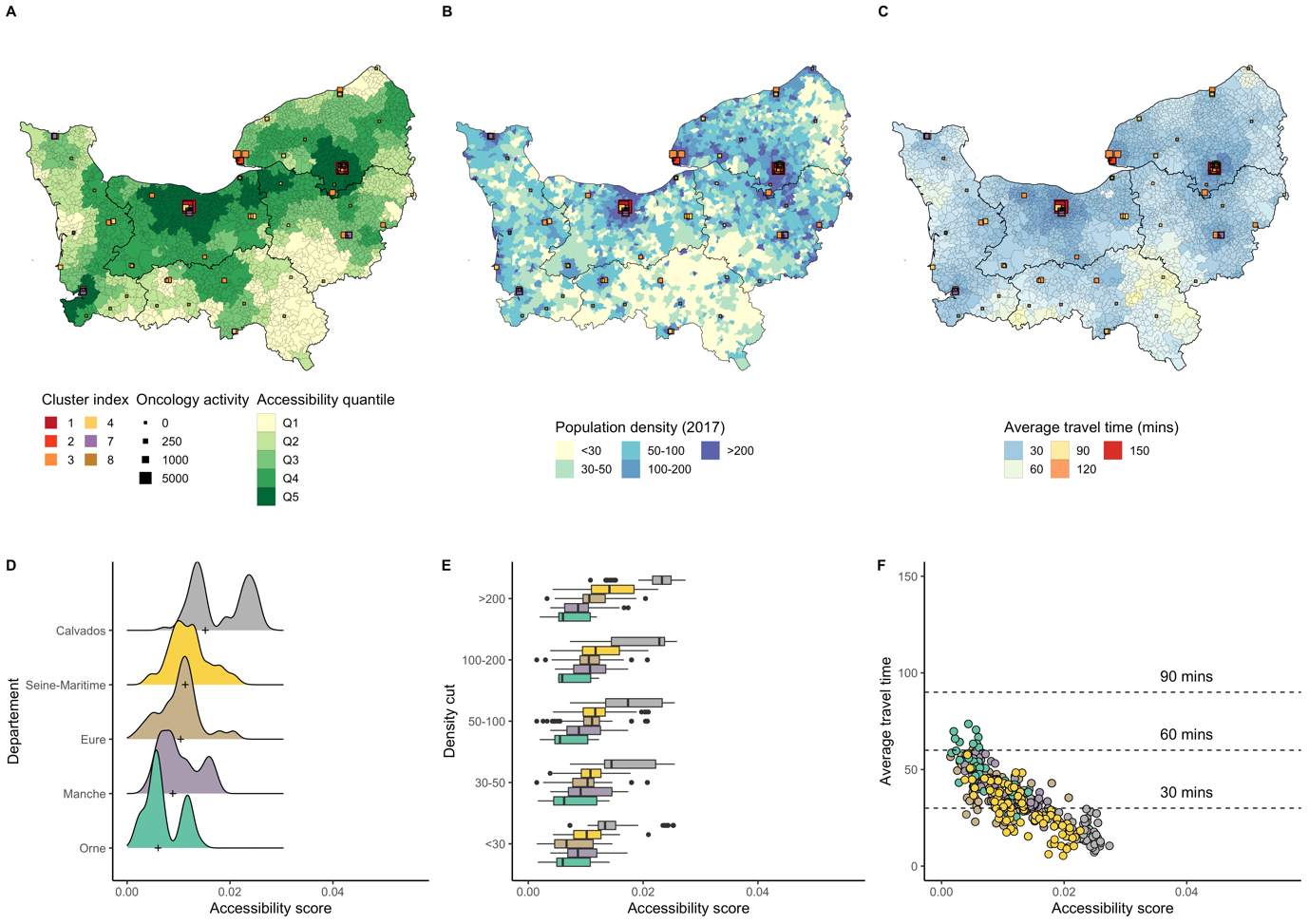

**Sup. Figure 24: Accessibility distribution in Normandie.**

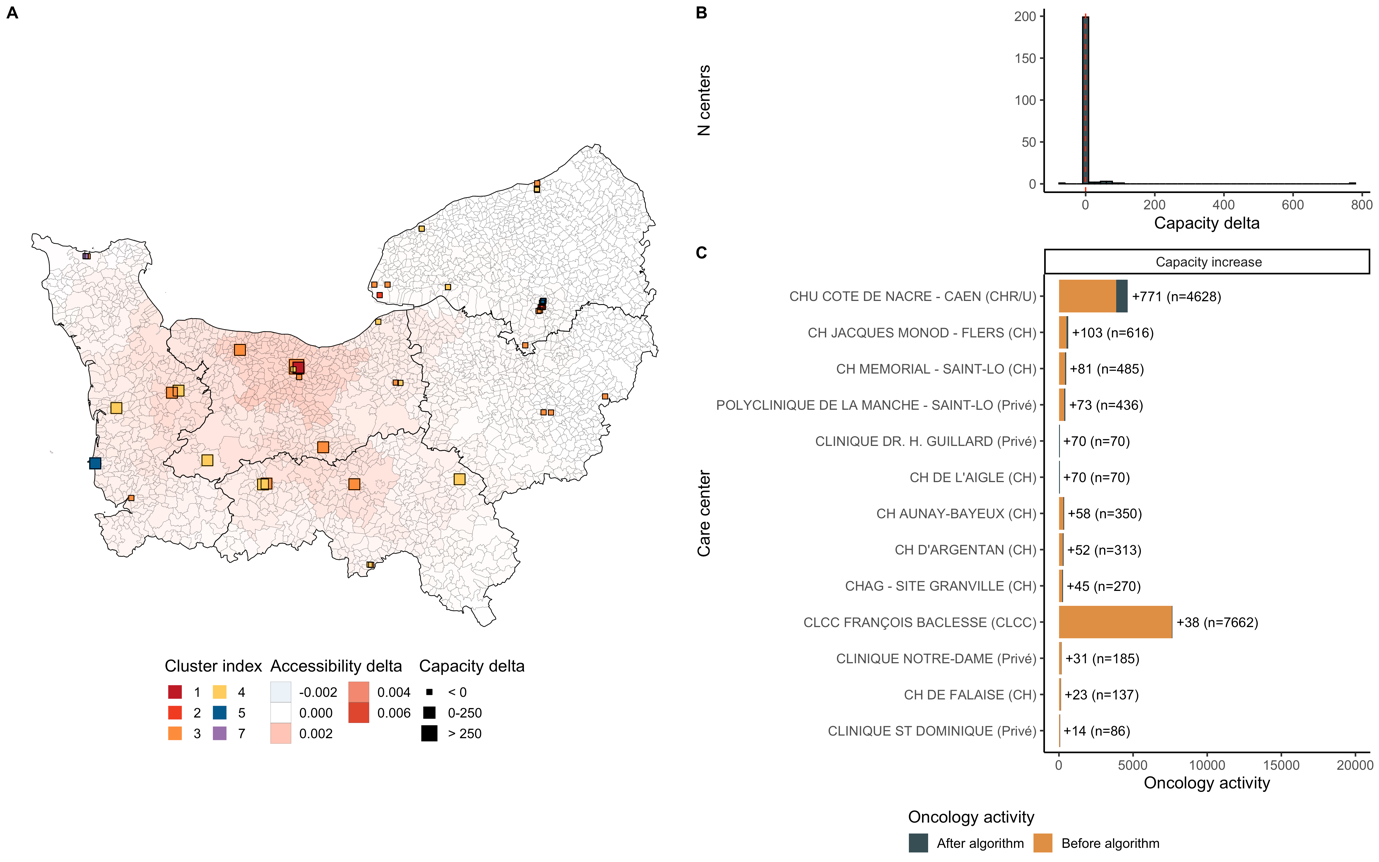

**Sup. Figure 25: Optimization results in Normandie.** Additional activity was 1,523. 15 centers grew and 0 decreased. Median accessibility before optimization was 0.0105 and 0.0106 after, corresponding to a 1% increase. Accessibility grew near Caen, Argentan, and St-Lo.

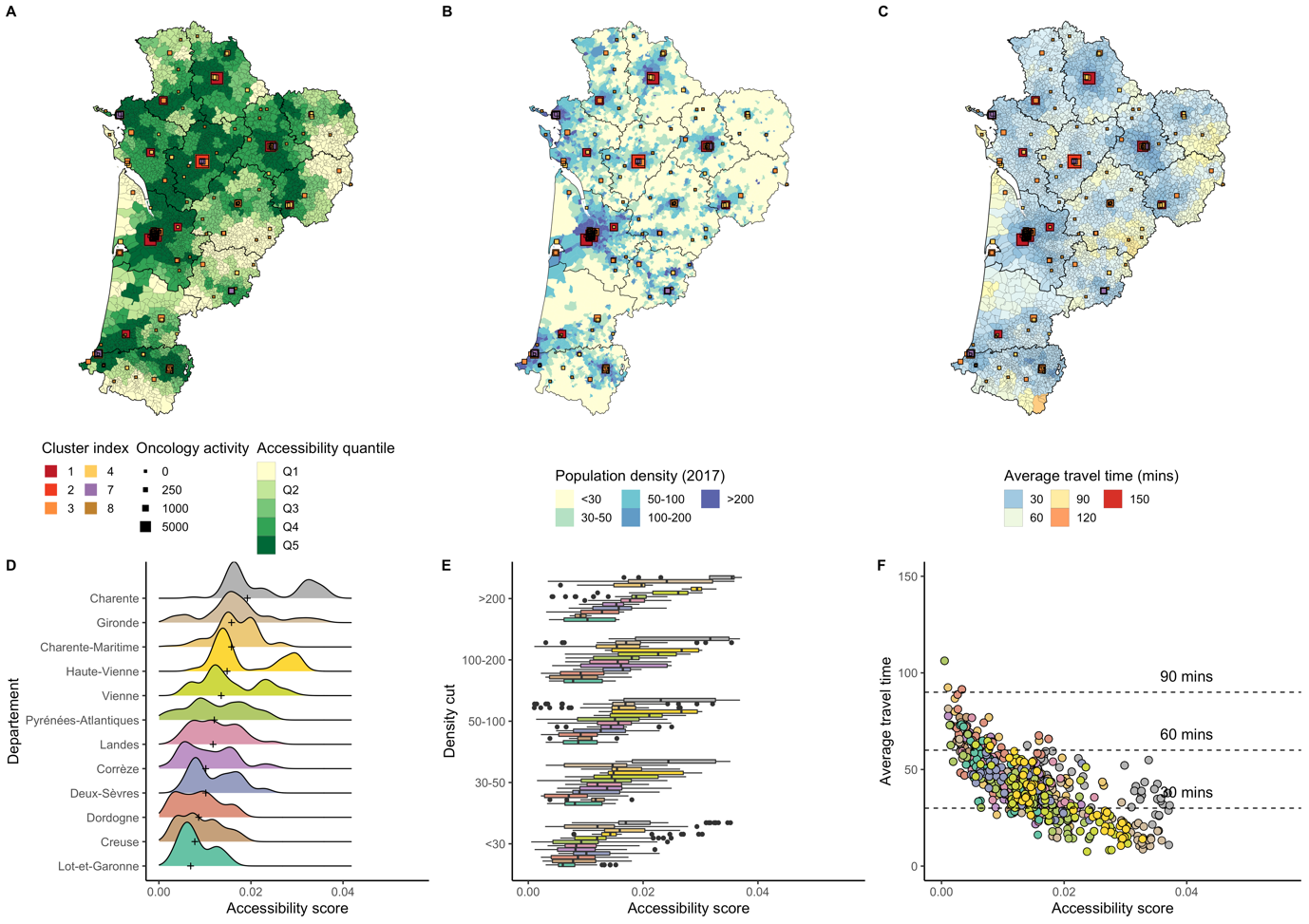

**Sup. Figure 26: Accessibility distribution in Nouvelle-Aquitaine.**

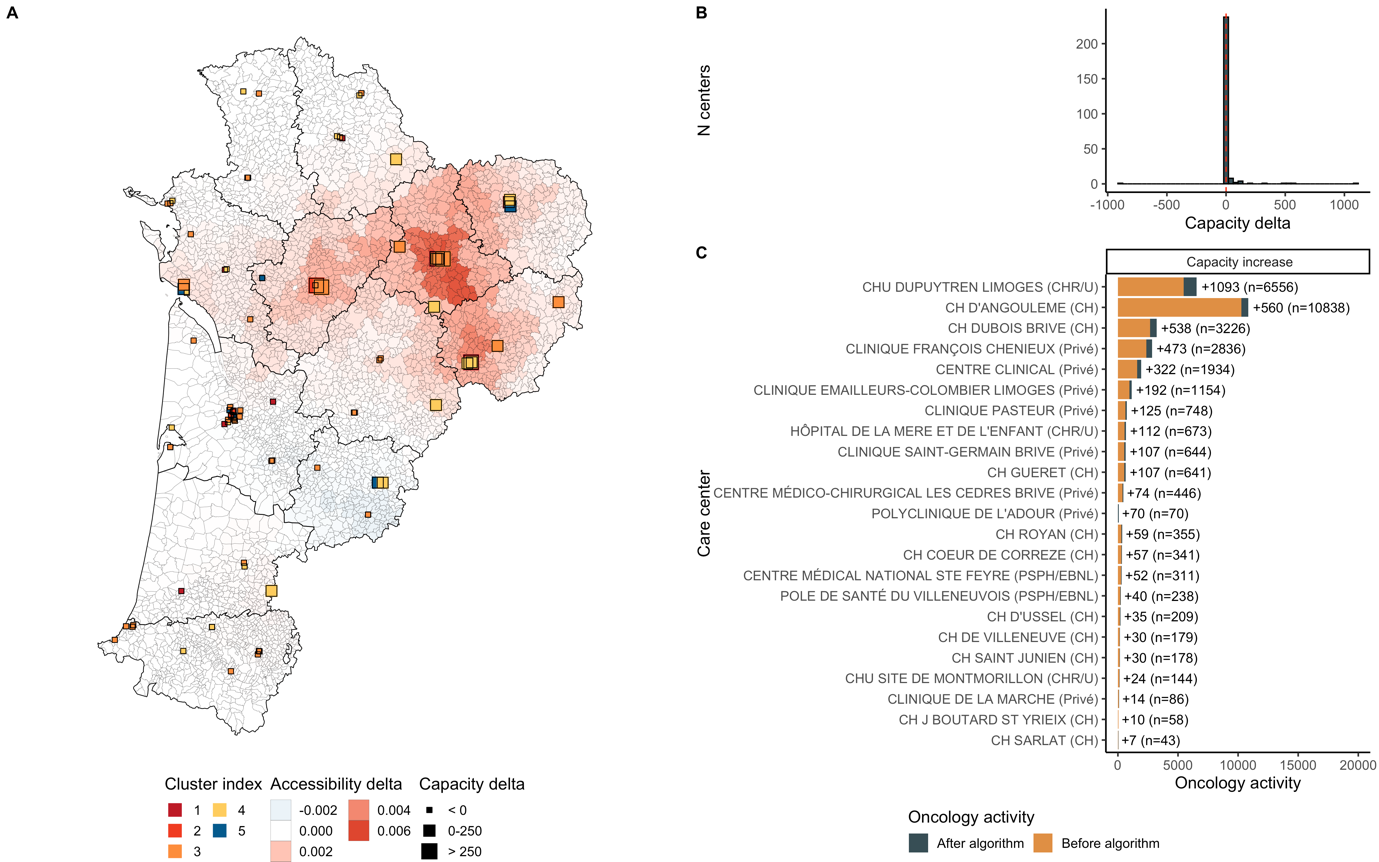

**Sup. Figure 27: Optimization results in Nouvelle-Aquitaine.** Additional activity was 3,445. 25 centers grew and 1 decreased. Median accessibility before optimization was 0.0117 and 0.0119 after, corresponding to a 1.5% increase. Accessibility grew around Limoges, Angouleme, and Brives-la-Gaillarde.

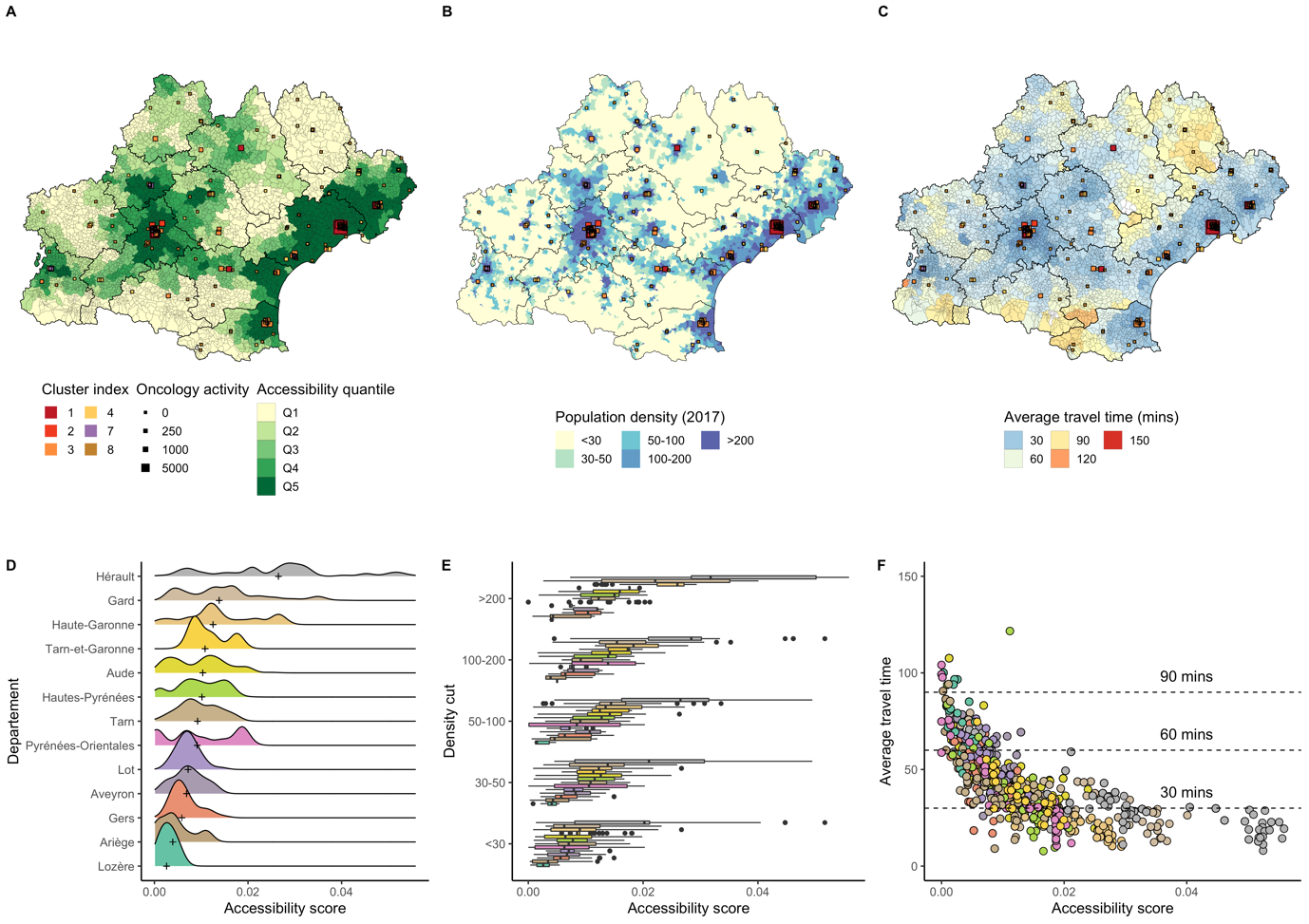

**Sup. Figure 28: Accessibility distribution in Occitanie.**

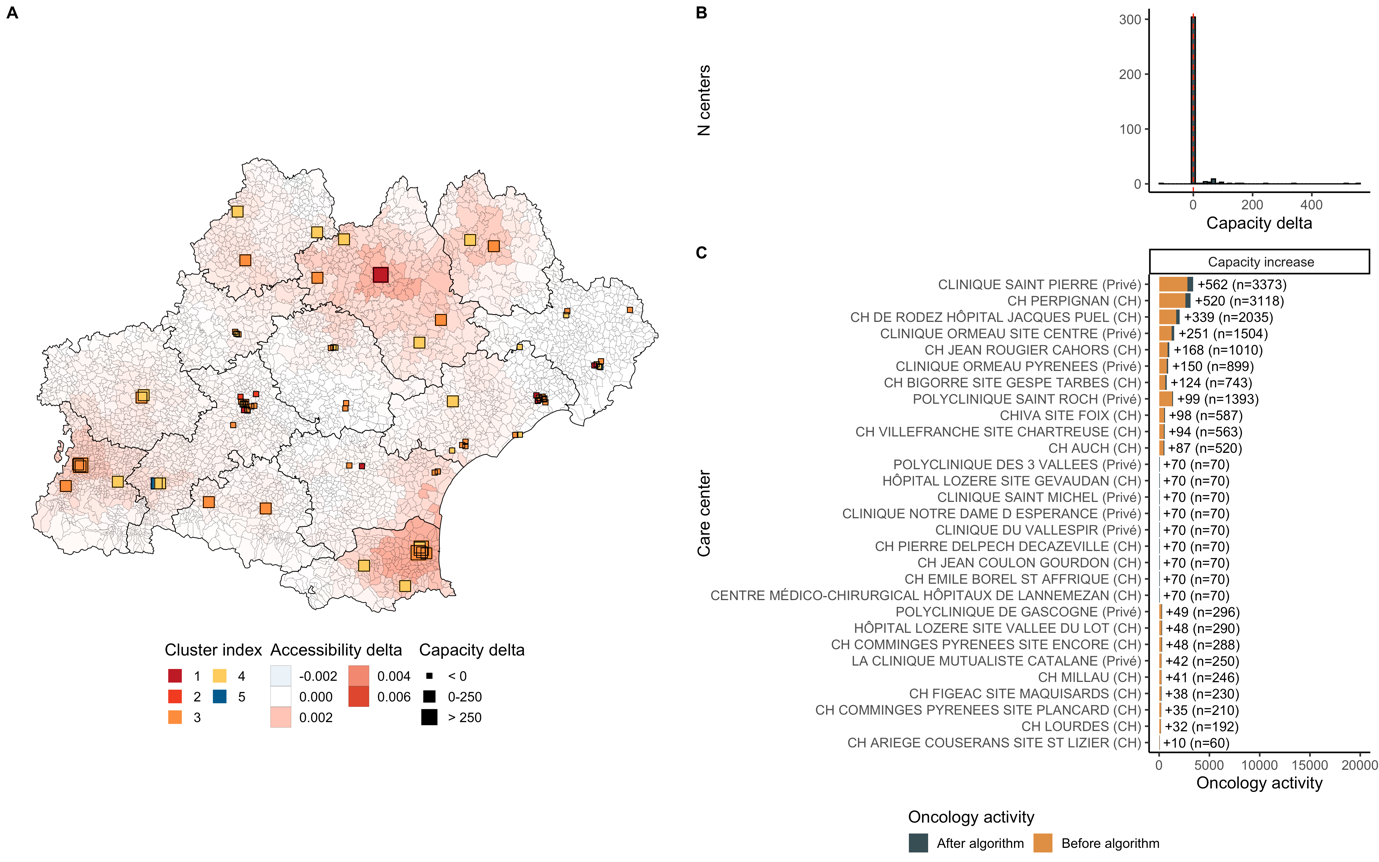

**Sup. Figure 29: Optimization results in Occitanie.** Additional activity was 3,652. 28 centers grew and 1 decreased. Median accessibility before optimization was 0.0087 and 0.0091 after, corresponding to a 4.7% increase. Accessibility grew around Perpignan, Rodez, Mende and Tarbes.

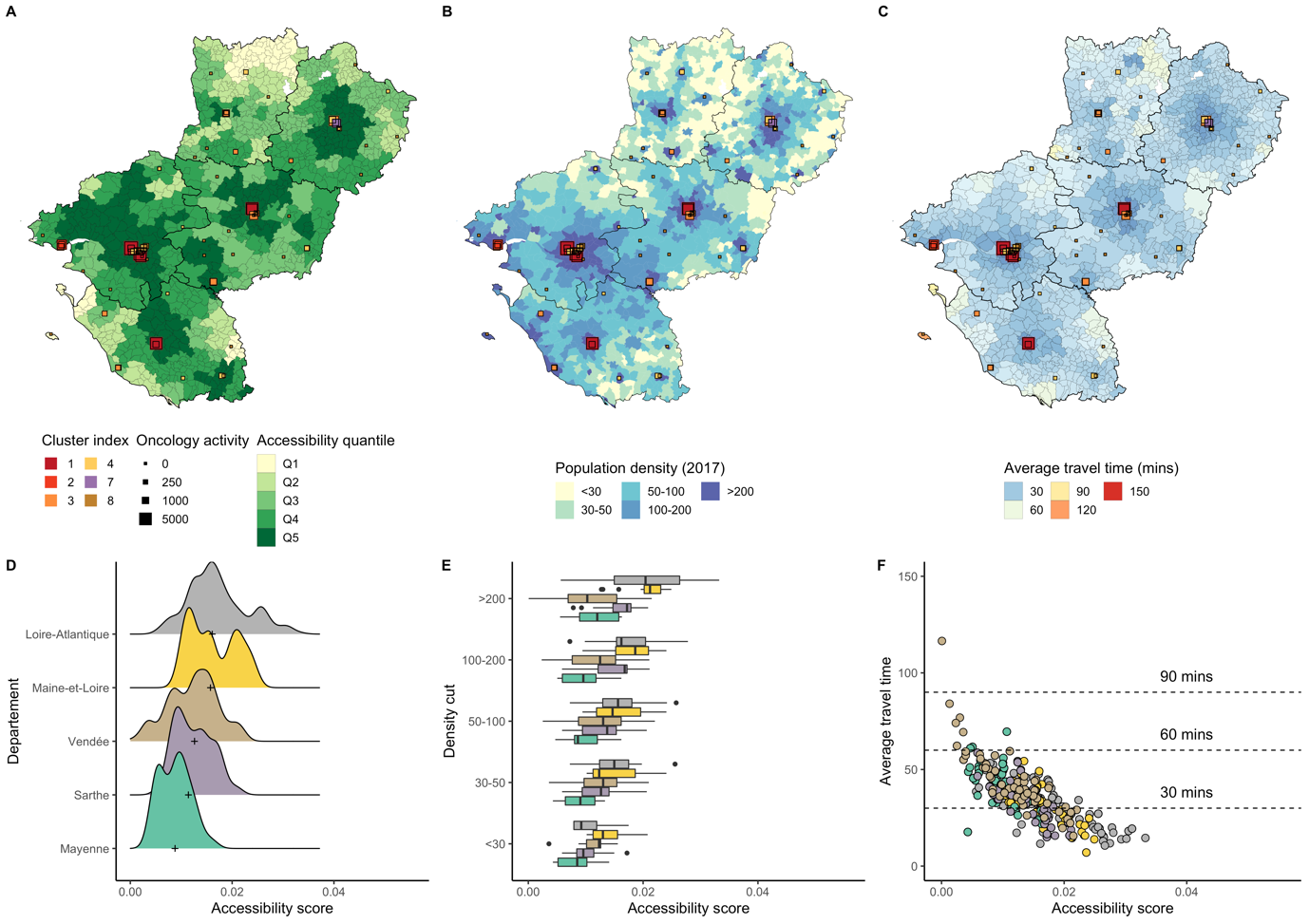

**Sup. Figure 30: Accessibility distribution in Pays-de-la-Loire.**

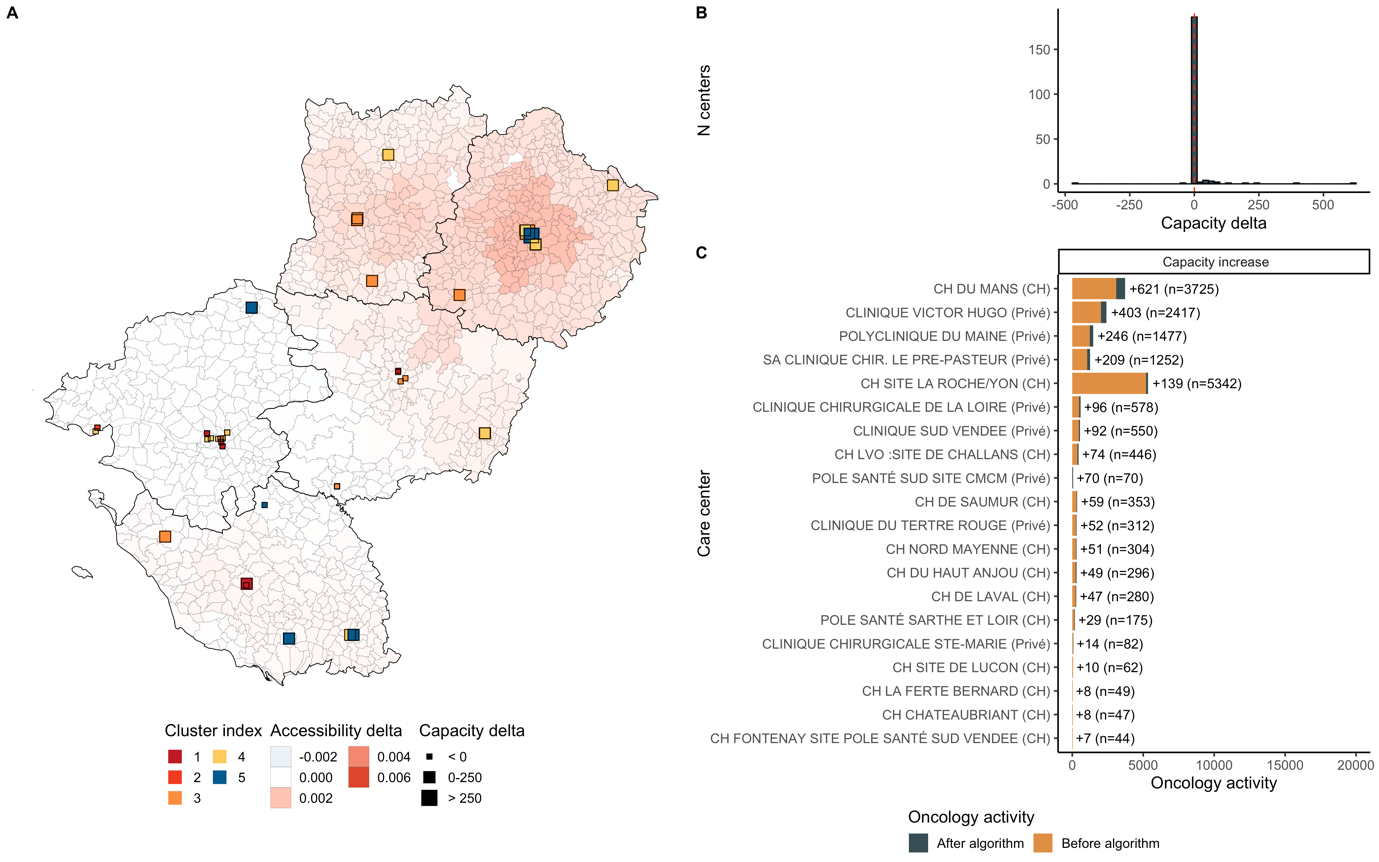

**Sup. Figure 31: Optimization results in Pays-de-la-Loire.** Additional activity was 1,890. 18 centers grew and 2 decreased. Median accessibility before optimization was 0.0118 and 0.0121 after, corresponding to a 2.4% increase. Accessibility grew near La Roche sur Yon, Angers and Le Mans.
